# Endpoint-aligned artificial intelligence for biopsy-sparing assessment of suspected basal cell carcinoma

**DOI:** 10.64898/2026.07.30.26359303

**Authors:** Zhaowei Chu, Qihao Duan, Yatao Guo, Wei Cao, Bingding Huang, Wanyu Zheng, Longfei Zhu, Roland Eils, Benjamin Wild, Songmei Geng, Lei Gu

## Abstract

Basal cell carcinoma (BCC) care follows a sequence of decisions from triage to pathological subtyping and depth assessment, and the information available changes at each step. To date, no artificial-intelligence (AI) tool using non-invasive inputs has been developed to support this full decision-making sequence. In this study, we developed a multi-endpoint AI framework matching non-invasive inputs to each decision point in 1,459 internal and 995 external patients. Triage macro-AUROC was 0.995 internally, 0.978 externally and 0.853 in a geographically distinct cohort, with risk stratification 0.943 and 0.899. For thickness, the highest-precision configuration used dermoscopy alone rather than all modalities (0.949 versus 0.881). Performance exceeded the 19-dermatologist mean on matched cases for every prespecified primary metric (all P ≤ 0.014). Local adaptation raised in-scope accuracy from 0.790 to 0.954 but shifted action-proxy routing toward the no-further-assessment classes for out-of-scope inputs, reducing sensitivity from 0.953 to 0.697. A validation-locked Mahalanobis gate enriched sensitivity among accepted cases to 0.775 at 0.791 coverage but only partially mitigated residual out-of-scope routing errors. These findings separate closed-set performance from scope control and support endpoint-specific validation of biopsy-sparing AI for BCC diagnosis and personalized treatment planning.

## Introduction

Basal cell carcinoma (BCC) is the most common keratinocyte carcinoma and a major contributor to the worldwide burden of nonmelanoma skin cancer ^1–3^. Early BCC can resemble benign lesions such as seborrhoeic keratosis (SK) and melanocytic naevus (MN), creating a common triage problem before histopathology is available ^4^. Routine BCC care, however, is more than a single malignant-versus-benign decision ^5^. Clinicians first decide whether a lesion is likely to be BCC or a mimic. Once BCC is suspected or confirmed, management also depends on anatomical site, histological growth pattern, tumour thickness and treatment goals ^6^.

Histopathology remains the reference standard for diagnosis, pathological subtype assessment and tumour thickness measurement. Biopsy is nonetheless invasive and costly, and access to dermatology or dermatopathology expertise may be limited. The accuracy of BCC pathological subtyping and thickness estimation can be compromised by incomplete tumour sampling and interobserver variability ^7,8^. This creates a practical need for non-invasive decision support that helps prioritize specialist assessment and biopsy while supporting later decisions in the care pathway.

The information available to a clinician prior to biopsy changes across the pathway. During remote triage and front-line assessment, the actionable information is dominated by visual inspection and brief clinical context, even though in-person examination can add tactile, dynamic and clinician-observed features ^9^. Specialist assessment may add dermoscopy, a modality with established diagnostic value for BCC recognition and wide accessibility in dermatology practice ^10,11^. A clinically useful AI framework should therefore be explicit about which endpoint it addresses, and which inputs are realistically available at that point in care.

Artificial intelligence (AI) has achieved strong performance in selected skin-cancer image-classification settings, and reader-facing studies have highlighted the importance of evaluating AI in relation to clinician decision-making rather than as an isolated algorithm ^9,12,13^. More recently, dermatology foundation models have provided reusable image representations for multimodal tasks ^14^. However, published nonmelanoma skin-cancer AI studies still primarily focus on imaging classification tasks, most commonly whole-slide imaging or dermoscopy, with fewer studies addressing linked management questions such as risk stratification or thickness support ^15–19^. Such systems are also trained on a closed label set, yet the lesions they meet in practice are not restricted to it. How a locally adapted model behaves on diagnoses outside its training classes, and whether selective prediction can bound that behaviour, is rarely reported. Here, we developed and externally validated a multi-endpoint BCC-centred AI framework using information obtained from non-invasive methods for accessible lesion triage, risk-aware stratification of BCC pathological subtypes and ≤2 mm tumour-thickness assessment. We evaluated endpoint-specific information sources and compared the primary AI configurations with a dermatologist panel on the same reader-study cases. We also examined how patient-grouped local adaptation changed in-scope performance and action routing for actinic keratosis (ACK), squamous cell carcinoma (SCC) and melanoma, evaluated four validation-locked selective-prediction scores, and inspected saliency maps to describe which image regions accompanied the model outputs at each endpoint.

## Results

### Cohorts and staged endpoint structure

The study included 1,459 patients in the XJTU cohort, 149 in the Baoji cohort and 846 in the Brazilian three-class cohort. The Brazilian cohort contained 1,066 lesions represented by 1,324 images. Demographics are reported at patient level, lesion distributions at lesion level and formal triage performance at image level (Table 1, Supplementary Table S1 and Supplementary Figure S2). XJTU provided model development and internal validation, Baoji provided external validation for all three endpoints, and Brazil provided a geographically distinct external triage cohort. The internal reader-study sets included 146 cases for lesion triage, 145 for risk-aware stratification and 24 for thickness prediction. Endpoint-specific model-only, reader-study and external analysis sets are shown in Figure 1A.

**Figure 1.**
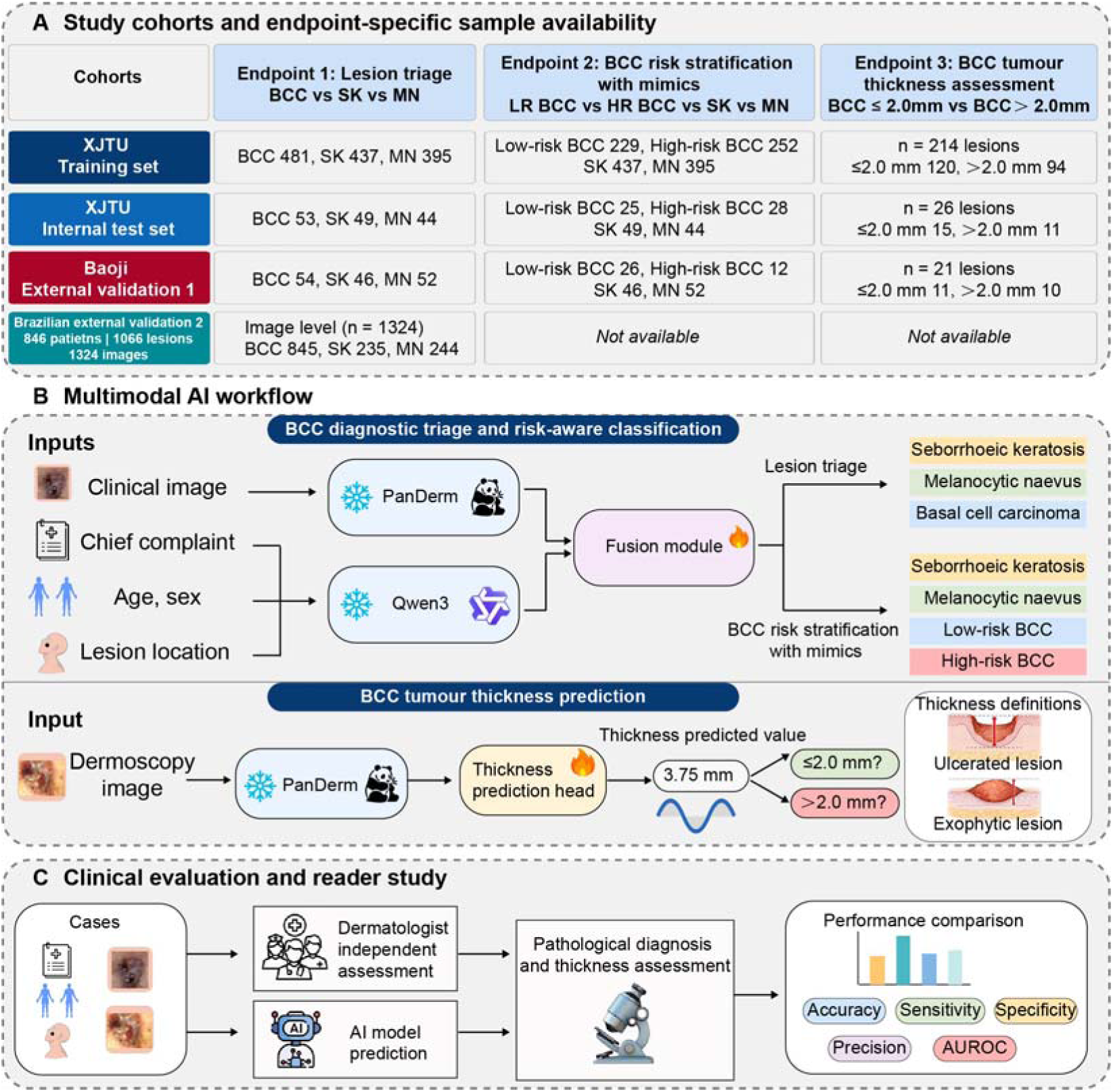
Study design and clinical validation. A, Cohorts and endpoint-specific analysis sets. Brazilian cohort totals are separated into patients, lesions and images. B, Endpoint-aligned inputs and model streams. Clinical photographs and structured context support triage and risk-aware stratification; dermoscopy supports the exploratory ≤2 mm thickness endpoint. C, Matched comparison with the shared 19-dermatologist reader panel. Counts in the artwork use SK, seborrhoeic keratosis; MN, melanocytic naevus; LR BCC and HR BCC, low-risk and high-risk BCC.

**Table 1.** Cohort characteristics with patient-, lesion- and image-level denominators.

| Characteristic | XJTU | Baoji | Brazil |
| --- | --- | --- | --- |
| **Cohort size** |  |  |  |
| Patients, n | 1,459 | 149 | 846 |
| Lesions, n | 1,459 | 152 | 1,066 |
| Clinical images in formal triage, n | 1,459 | 152 | 1,324 |
| **Patient-level demographics** |  |  |  |
| Age, years, mean (SD) | 52.4 (17.8) | 52.8 (17.3) | 57.4 (17.5) |
| Female, n (%) | 905 (62.0) | 89 (59.7) | 297 (35.1) |
| Male, n (%) | 554 (38.0) | 60 (40.3) | 278 (32.9) |
| Sex not reported, n (%) | 0 | 0 | 271 (32.0) |
| **Lesion-level anatomical site** |  |  |  |
| Head and face, n (%) | 982 (67.3) | 96 (63.2) | 545 (51.1) |
| Trunk, n (%) | 362 (24.8) | 45 (29.6) | 327 (30.7) |
| Extremities, n (%) | 115 (7.9) | 11 (7.2) | 194 (18.2) |
| **Lesion-level triage diagnosis** |  |  |  |
| BCC, n (%) | 534 (36.6) | 54 (35.5) | 652 (61.2) |
| Seborrhoeic keratosis, n (%) | 486 (33.3) | 46 (30.3) | 198 (18.6) |
| Melanocytic naevus, n (%) | 439 (30.1) | 52 (34.2) | 216 (20.3) |
| **BCC histopathological risk subgroup** |  |  |  |
| Low-risk BCC, n (% of BCC) | 254 (47.6) | 26 (48.1) | — |
| High-risk BCC, n (% of BCC) | 280 (52.4) | 12 (22.2) | — |
| Risk subtype not annotated, n (% of BCC) | 0 | 16 (29.6) | — |
| Dermoscopy available, n/BCC (%) | 240/534 (44.9) | 31/54 (57.4) | — |

Brazilian cohort totals are 846 patients, 1,066 lesions and 1,324 images. Age and sex are patient-level; site and diagnosis are lesion-level; formal Brazilian triage performance is image-level. For 36 Brazilian patients with inconsistent age entries across image records, the median recorded age was used. Head and face comprised FACE, NOSE, NECK, EAR, SCALP and LIP; trunk comprised BACK, CHEST and ABDOMEN; extremities comprised FOREARM, ARM, THIGH, HAND and FOOT. XJTU contributed one selected lesion and clinical image per patient. In Baoji, three patients each contributed two lesions at the same visit, so 149 patients provided 152 lesions and 152 images.

### Pre-biopsy lesion triage across validation cohorts

For pre-biopsy lesion triage, BCC versus seborrhoeic keratosis versus melanocytic naevus, the primary visual-contextual model achieved a macro-averaged area under the receiver operating characteristic curve (AUROC) of 0.995 (0.986-0.999) internally, 0.978 (0.963-0.990) in the Baoji cohort and 0.853 (0.838-0.868) in the Brazilian cohort (Table 2). Accuracy was 0.944 (0.916-0.967) internally, 0.836 (0.791-0.878) in the Baoji cohort and 0.784 (0.772-0.797) in the Brazilian cohort. Macro-averaged precision and specificity remained above 0.86 and 0.91 in the Baoji cohort, respectively (Figure 2). Extended external validation rows and class-specific AUROC estimates are provided in Supplementary Table S3 and Supplementary Figure S3.

**Figure 2.**
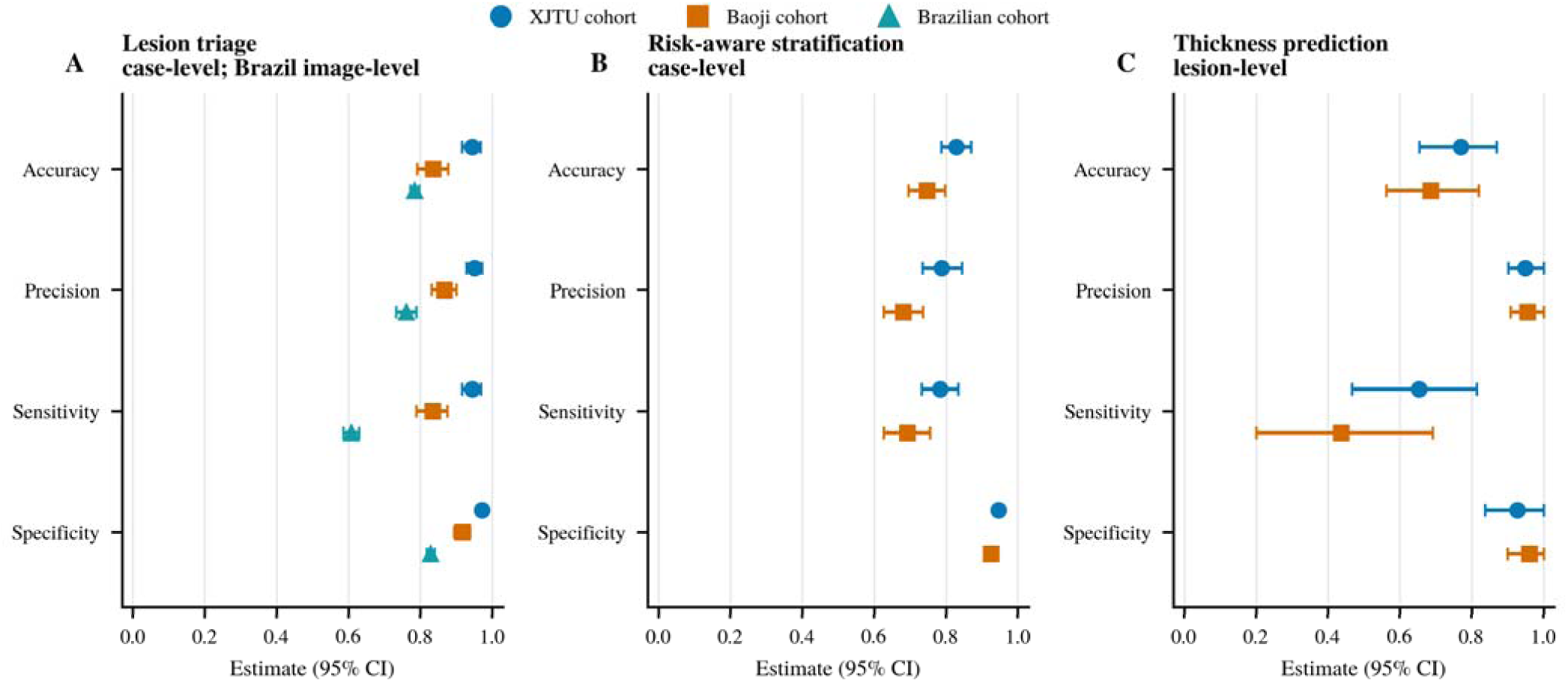
Primary endpoint performance. A, Three-class lesion triage. B, Four-class risk-aware stratification. C, Exploratory ≤2 mm thickness prediction. Points are five-seed means and horizontal intervals are 95% case-bootstrap confidence intervals. Cohorts are encoded by both colour and marker shape. Brazilian triage is image-level; the Chinese classification analyses are case-level and thickness is lesion-level.

**Table 2.** Primary model performance and matched dermatologist comparisons.

| Analysis set (unit) | Comparator | Accuracy | Precision | Sensitivity | Specificity | AUROC | P value |
| --- | --- | --- | --- | --- | --- | --- | --- |
| **Lesion triage** |  |  |  |  |  |  |  |
| XJTU reader set (146 cases) | AI | 0.944 (0.914-0.968) | 0.951 (0.928-0.971) | 0.944 (0.914-0.969) | 0.972 (0.957-0.984) | 0.995 (0.986-0.999) | <0.001 |
| XJTU reader set (146 cases) | 19 dermatologists | 0.817 (0.783-0.849) | 0.831 (0.802-0.863) | 0.818 (0.785-0.849) | 0.909 (0.892-0.925) | — | — |
| Baoji external (152 cases) | AI | 0.836 (0.791-0.878) | 0.867 (0.831-0.901) | 0.834 (0.789-0.876) | 0.917 (0.894-0.938) | 0.978 (0.963-0.990) | — |
| Brazil external (1,324 images) | AI | 0.784 (0.772-0.797) | 0.761 (0.733-0.789) | 0.608 (0.586-0.630) | 0.828 (0.817-0.839) | 0.853 (0.838-0.868) | — |
| **Risk-aware stratification** |  |  |  |  |  |  |  |
| XJTU reader set (145 cases) | AI | 0.829 (0.788-0.869) | 0.787 (0.734-0.842) | 0.782 (0.730-0.833) | 0.946 (0.932-0.959) | 0.942 (0.920-0.963) | <0.001 |
| XJTU reader set (145 cases) | 19 dermatologists | 0.692 (0.654-0.725) | 0.660 (0.625-0.696) | 0.658 (0.619-0.692) | 0.899 (0.887-0.910) | — | — |
| Baoji external (136 cases) | AI | 0.747 (0.696-0.797) | 0.680 (0.627-0.736) | 0.693 (0.627-0.756) | 0.925 (0.910-0.940) | 0.899 (0.863-0.932) | — |
| **≤2 mm thickness** |  |  |  |  |  |  |  |
| XJTU model-only (26 lesions) | AI | 0.769 (0.654-0.869) | 0.949 (0.902-1.000) | 0.653 (0.467-0.813) | 0.927 (0.836-1.000) | — | — |
| XJTU reader set (24 lesions) | AI | 0.758 (0.650-0.867) | 0.960 (0.913-1.000) | 0.653 (0.493-0.827) | 0.933 (0.844-1.000) | — | 0.014 |
| XJTU reader set (24 lesions) | 19 dermatologists | 0.634 (0.531-0.730) | 0.852 (0.743-0.948) | 0.530 (0.382-0.667) | 0.807 (0.667-0.924) | — | — |
| Baoji external (21 lesions) | AI | 0.686 (0.562-0.819) | 0.956 (0.908-1.000) | 0.436 (0.200-0.691) | 0.960 (0.900-1.000) | — | — |

Values are five-seed means (95% CI). For the classification endpoints, precision, sensitivity and specificity are macro averages; for the ≤2 mm thickness endpoint they are binary values for the ≤2 mm positive class. P values use the prespecified reader-comparison endpoint: macro-F1 for classification and precision for thickness, with paired stratified case-bootstrap comparisons of AI with the 19-reader mean. XJTU risk model-only AUROC was 0.943 (0.921-0.964; n=146); 0.942 is the matched reader-set estimate (n=145).

Among the two external cohorts, the addition of structured text produced the larger gain in Baoji. The clinical image-only model achieved macro-AUROC 0.899 (0.865-0.931) and accuracy 0.696 (0.645-0.747), compared with 0.978 (0.963-0.990) and 0.836 (0.791-0.878), respectively, for clinical image plus text. In the Brazilian cohort, structured text produced a smaller increase in macro-AUROC, from 0.837 (0.824-0.850) with clinical images alone to 0.853 (0.838-0.868) with clinical images and text (Supplementary Table S3).

### BCC risk stratification while preserving common mimics

The BCC risk stratification endpoint retained the two non-BCC mimics, seborrhoeic keratosis and melanocytic naevus, while separating BCC into low- and high-risk histopathological groups. This task was more difficult than triage but retained discrimination well above chance, with macro-AUROC 0.943 (0.921-0.964) internally and 0.899 (0.863-0.932) in the Baoji cohort (Table 2). Accuracy was 0.829 (0.786-0.870) internally and 0.747 (0.696-0.797) externally (Figure 2).

Among pathology-confirmed BCC cases, we also evaluated high-risk versus low-risk subtypes as conditional binary endpoints (Supplementary Figure S5). The primary risk-stratification result remained the four-class endpoint because it assessed BCC risk groups while retaining SK and MN in the same output space.

Because the 4-class endpoint separated BCC into low- and high-risk subclasses while retaining SK and MN, we tested whether broader three-class discrimination was preserved after BCC subclassification. On 136 matched Baoji cases, the dedicated 3-class model achieved accuracy 0.825 (95% CI 0.775-0.872), macro-averaged sensitivity 0.839 (0.791-0.882) and macro-averaged specificity 0.917 (0.893-0.939). After collapsing the trained 4-class model’s low- and high-risk BCC outputs, the corresponding estimates were 0.882 (0.838-0.922), 0.888 (0.844-0.926) and 0.944 (0.923-0.963), with overlapping intervals (Supplementary Figure S9 and Supplementary Note S6). In this matched external comparison, the broader BCC/SK/MN discrimination was retained when the BCC subclasses were recombined.

### Thickness prediction after dermoscopy-based assessment

For the ≤2 mm thickness endpoint, we prioritized precision. Dermoscopy-only was selected as the primary model after specialist-stage lesion assessment. Using seed-specific validation-derived F0.5 thresholds, the model achieved precision 0.949 (0.902-1.000) in the XJTU model-only internal test set (n=26) and 0.956 (0.908-1.000) in Baoji (n=21). The corresponding sensitivities were 0.653 (0.467-0.813) and 0.436 (0.200-0.691), and specificities were 0.927 (0.836-1.000) and 0.960 (0.900-1.000), respectively (Figure 2C and Supplementary Table S4). This operating point therefore prioritized precision over sensitivity.

Thickness prediction illustrated that more inputs did not necessarily produce a better operating model. Under the validation-derived F0.5 policy, clinical image plus dermoscopy plus text reached precision 0.881 (0.781-1.000) in XJTU and 0.858 (0.646-1.000) in Baoji, whereas dermoscopy-only reached 0.949 (0.902-1.000) and 0.956 (0.908-1.000), respectively (Supplementary Table S4). Dermoscopy-only therefore gave the highest precision of the modality combinations evaluated under this policy. Modality and threshold-policy comparisons are shown in Supplementary Tables S4 and S5 and Supplementary Figures S6 and S7.

### Comparison with dermatologists

In the internal reader-study analysis set, AI seed-mean estimates were higher than the mean of the 19-reader panel for the prespecified primary metric of each endpoint in the same cases (Figure 3). For 3-class diagnosis, AI macro-F1 was 0.945 (0.915-0.969) versus 0.813 (0.779-0.845) for dermatologists (P < 0.001). For 4-class diagnosis, AI macro-F1 was 0.778 (0.724-0.828) versus 0.642 (0.602-0.675) (P < 0.001). For thickness prediction, AI precision was 0.960 (0.913-1.000) versus 0.852 (0.743-0.948) (P = 0.014). These comparisons benchmark the model against the reader panel under matched case conditions.

**Figure 3.**
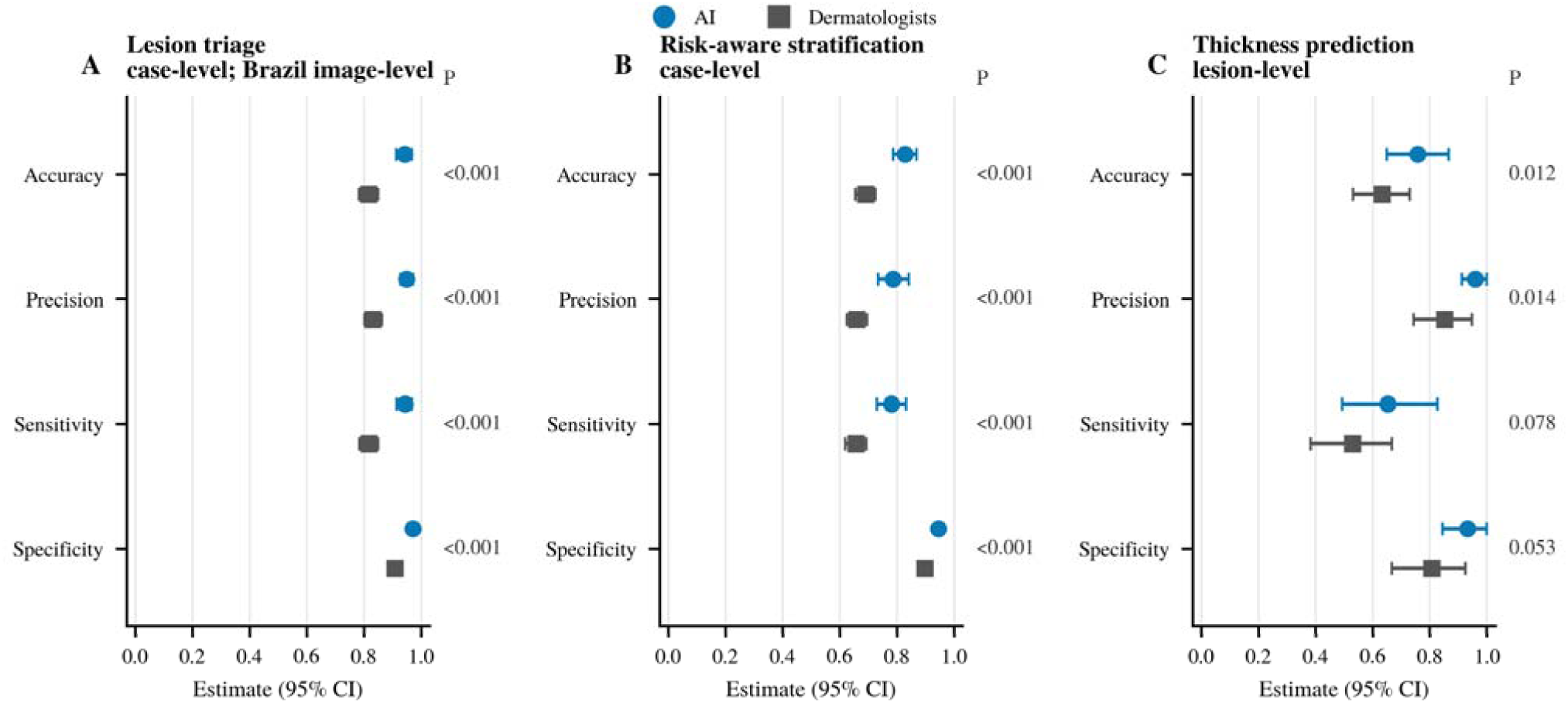
AI and dermatologist performance on matched internal reader-study cases. A, Lesion triage (146 cases). B, Risk-aware stratification (145 cases). C, ≤2 mm thickness prediction (24 lesions). Points show the AI five-seed mean or mean across 19 dermatologists; intervals are paired case-bootstrap 95% confidence intervals. The right-hand column of each panel prints the paired stratified case-bootstrap P value for that metric; the prespecified primary metric of each endpoint is macro-F1 (A, B) and precision (C), and the remaining values are exploratory and unadjusted. An em dash marks a metric for which no reader comparison was defined.

#### Local adaptation improves in-scope discrimination but shifts out-of-scope routing

Given the lower external-transfer performance of the XJTU-trained model in Brazil, we first evaluated patient-grouped local adaptation using a fixed patient-disjoint Brazilian test set. In the image-level adaptation-scaling experiment, clinical image plus text accuracy increased from 0.812 (0.768-0.853) without local adaptation to 0.861 (0.820-0.898) with a training pool targeting 64 Brazilian images and 0.971 (0.947-0.991) with 1,192 images. The corresponding macro-AUROC values were 0.905 (0.859-0.942), 0.942 (0.908-0.970) and 0.998 (0.995-1.000), respectively. Training pools and the fixed test set, which contained 132 images from 78 lesions in 27 patients, were patient-disjoint (Supplementary Figure S4; Supplementary Note S5).

We next examined whether this in-scope improvement was accompanied by safer routing of diagnoses outside the BCC/SK/MN label set. After aggregating probabilities at lesion level and excluding every patient used for adaptation training, the six-diagnosis action-proxy cohort comprised 898 images from 728 lesions in 554 patients, including 700 lesions assigned to the further-assessment reference group and 28 assigned to the non-action group. Accuracy on the fixed 78-lesion in-scope test set increased from 0.790 (0.664-0.887) for the unadapted XJTU-trained model to 0.954 (0.894-0.997) for the Brazil-adapted model (Figure 4A). In the six-diagnosis cohort, action-proxy sensitivity decreased from 0.953 (0.940-0.964) to 0.697 (0.668-0.726), whereas specificity increased from 0.579 (0.415-0.729) to 0.979 (0.931-1.000), balanced accuracy increased from 0.766 to 0.838. However, the change was diagnosis specific (Figure 4C and Supplementary Figure S10), reflecting a shift in the sensitivity-specificity operating point rather than a uniform improvement in action routing (Figure 4B). Thus, local adaptation improved in-scope performance but shifted out-of-scope routing toward the no-further-assessment classes, indicating that these two objectives are related but distinct that require separate validation.

**Figure 4.**
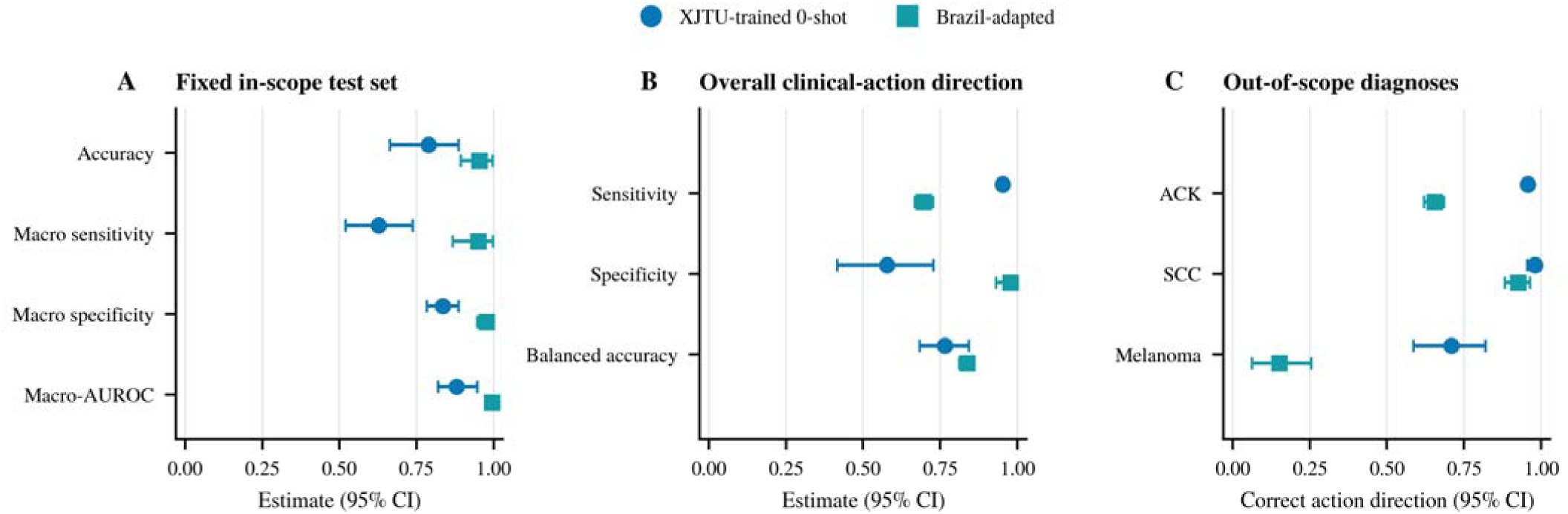
Local adaptation improves in-scope discrimination but shifts out-of-scope action-proxy routing. A, Lesion-level three-class accuracy, macro sensitivity, macro specificity and macro-AUROC on the fixed in-scope test set (78 lesions from 27 patients) for the unadapted XJTU-trained and Brazil-adapted models. B, Action-proxy sensitivity, specificity and balanced accuracy in the combined six-diagnosis cohort after exclusion of all patients used for adaptation training. C, Proportion routed to further assessment for ACK (515 lesions from 428 patients), SCC (106 lesions from 97 patients) and melanoma (29 lesions from 29 patients). Panel B uses 728 lesions from 554 patients. Macro sensitivity and macro specificity are averaged over the three in-scope classes. Balanced accuracy is reported for the action proxy because the further-assessment and non-action reference groups are strongly unbalanced (700 versus 28 lesions). Points are five-seed means and intervals use 2,000 patient-cluster bootstrap replicates. The action proxy was derived from the three-class outputs, with BCC representing further assessment and SK/MN the non-action route; it does not represent observed clinical management decisions.

#### Validation-locked abstention enriches accepted-set performance but leaves residual out-of-scope risk

We next evaluated whether validation-locked selective-prediction gates could defer potentially unreliable predictions. For each model, seed and score, thresholds for maximum softmax probability (MSP), entropy, energy and Mahalanobis distance were fixed at the 95th percentile of in-scope validation lesion scores. Prediction correctness was not used to select the thresholds, and no out-of-scope evaluation lesion was used for calibration.

In the Brazil-adapted model, all four gates increased accepted-only action-proxy sensitivity relative to the ungated value of 0.697, but at reduced coverage (Figure 5 and Supplementary Table S6b). These estimates quantify enrichment among retained lesions and should not be interpreted as whole-cohort performance improvements. Abstention was also diagnosis dependent: the four gates deferred 2.1-5.9% of in-scope lesions and intercepted only 12.1-18.3% of melanoma predictions that would otherwise have been routed to SK/MN (Supplementary Figure S12).

**Figure 5.**
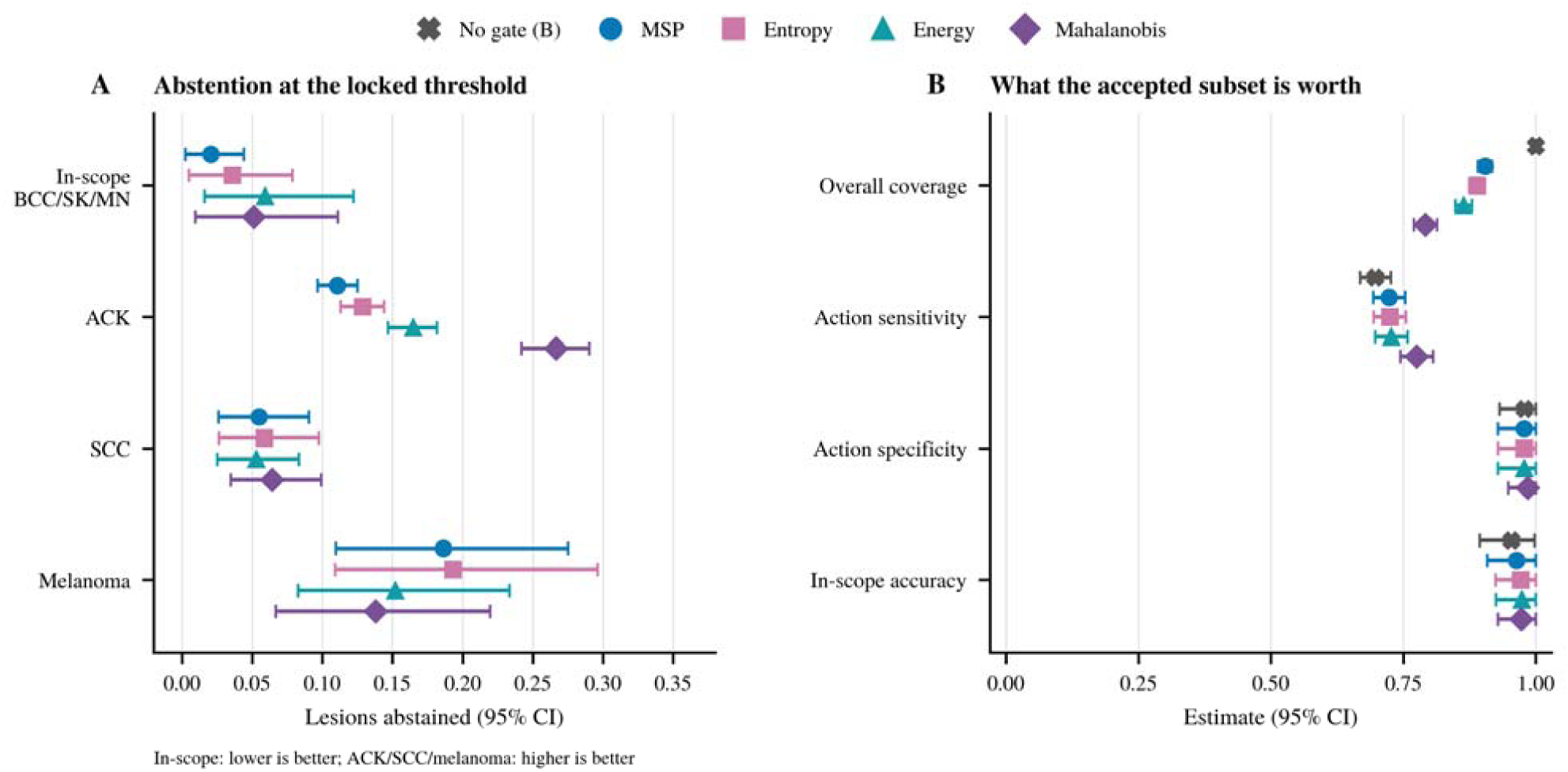
Validation-locked selective prediction enriches accepted-set performance but leaves residual out-of-scope risk. All panels show the Brazil-adapted model; Supplementary Figure S11 reports all four gates in both models. A, Diagnosis-specific proportions of in-scope, ACK, SCC and melanoma lesions abstained by MSP, entropy, energy and Mahalanobis at validation-locked thresholds. B, Overall coverage, accepted-only action-proxy sensitivity and specificity in the combined action cohort (728 lesions from 554 patients), and accepted-only accuracy among the 78 in-scope lesions from 27 patients. The ungated result is shown on every row of B as a reference; abstention without a gate is zero by definition, so no reference is shown in A. Points are lesion-level five-seed means and intervals use 2,000 patient-cluster bootstrap replicates. Accepted-only metrics use non-abstained lesions as the denominator and quantify enrichment among retained lesions rather than whole-cohort performance improvement; coverage is the proportion of all lesions retained. Mahalanobis was carried forward for exploratory detailed reporting only after all four scores had been compared, and the gates were not formally ranked. Diagnosis-specific residual accepted SK/MN routing is reported for both models in Supplementary Figure S12.

Mahalanobis yielded the numerically highest accepted-only sensitivity in both models and was carried forward as an exploratory post-comparison example; the four gates were not formally ranked. At its locked threshold, accepted-only action-proxy sensitivity for the Brazil-adapted model was 0.775 (0.744-0.806) at overall coverage of 0.791 (0.770-0.813). However, 0.745 (0.600-0.871) of melanomas remained accepted and routed to SK/naevus. Selective prediction therefore identified a higher-sensitivity accepted subset but did not provide reliable control of residual out-of-scope routing (Figure 5; Supplementary Figures S11-S12; Supplementary Tables S6a and S6b).

### Interpretation

Gradient-weighted class activation mapping (Grad-CAM) heatmaps were generated to show image regions associated with the model outputs (Supplementary Note S4). In the selected three-class examples, the highest saliency was often near the lesion margin. In the risk-stratification examples, saliency included lesion shape, contour and surrounding skin, whereas the thickness examples showed saliency across the lesion surface and surrounding skin (Supplementary Figure S8). These examples are descriptive and were not used for model selection or biological interpretation.

## Discussion

In this study, we developed a multi-endpoint BCC-centred AI framework using pre-biopsy information for three staged questions: lesion triage among BCC and common mimics, histopathology-informed risk stratification while retaining those mimics, and exploratory ≤2 mm tumour-thickness assessment. To our knowledge, this is the first BCC-centred framework to span these three staged endpoints and, in the same cohorts, to report how patient-grouped local adaptation changes action routing for diagnoses outside the trained label set under validation-locked abstention thresholds. Triage macro-AUROC was above 0.85 in all three cohorts, and risk-stratification macro-AUROC was 0.943 internally and 0.899 in Baoji. Dermoscopy-only thickness prediction achieved precision 0.949 internally and 0.956 externally. AI estimates were higher than the mean of 19 dermatologists on the same reader-study cases. The adaptation and selective-prediction analyses further showed that closed-set improvement, out-of-scope action routing and performance in an accepted subset are related but distinct properties. Previous nonmelanoma skin-cancer AI work has focused mainly on isolated image-classification tasks, most commonly dermoscopy ^20^, rather than a linked sequence of triage, risk grouping and thickness assessment. Our study examined pre-biopsy information across three related clinical questions and matched the input set to the information available at each point. Adding structured text to clinical images improved external triage in Baoji (macro-AUROC 0.978 versus 0.899 for image only) and produced a smaller improvement in Brazil (0.853 versus 0.837). Conversely, adding every modality did not improve the thickness endpoint. The Brazil comparison also combined differences in imaging, annotation, population and available metadata, and these effects cannot be separated in the present data.

The BCC risk-stratification endpoint reframed triage as a single classifier that retained two common mimics while separating BCC into low- and high-risk histopathological groups. Performance decreased from three-class triage to the native four-class task, as expected for a pathology-defined distinction that is challenging even on whole-slide histopathology ^21^, but the matched collapse analysis was consistent with preserved BCC/SK/MN discrimination after BCC subclassification. This supports risk-aware prioritisation within the staged framework while retaining the common mimics in the same output space.

The tumour-thickness endpoint illustrated that adding every available modality did not yield the strongest model. Dermoscopy makes vascular patterns, ulceration and lesion structure more visible than clinical photographs, whereas the structured text fields contained little direct information about thickness. Competition between modalities can also cause multimodal models to underuse weaker inputs ^22^. Reflectance confocal microscopy and optical coherence tomography can provide additional non-invasive thickness information but require specialised equipment and local expertise ^23,24^. The precision-prioritized dermoscopy model therefore offers a rule-in signal for further study. External sensitivity was 0.436, so a negative prediction carries little information, and the 26-lesion internal and 21-lesion external sets keep this endpoint exploratory and unsuitable for treatment selection.

On the shared reader-study cases, AI estimates exceeded the mean of the 19-dermatologist panel for the primary metric of each endpoint. Because both were evaluated on the same cases, these comparisons provide a direct benchmark for the three endpoint-specific configurations. The study was not designed to measure AI-assisted clinical performance, which remains a separate prospective question.

Patient-grouped local adaptation substantially improved discrimination within the Brazilian BCC/SK/MN label space but shifted the model’s action-proxy operating profile for diagnoses outside that space. The high action sensitivity of the unadapted model largely reflected its tendency to assign BCC broadly: it predicted BCC for 93.2% of lesions in a cohort in which 96.2% were assigned to the further-assessment reference group. Its melanoma referral rate should therefore not be interpreted as evidence that the model recognised melanoma. After adaptation, improved separation among BCC, SK and MN increased action specificity but was accompanied by more melanomas being mapped to the two non-action classes. This pattern is consistent with sharper closed-set discrimination rather than loss of a previously demonstrated melanoma-detection capability. Local adaptation within a defined label space and safe behaviour beyond that label space should therefore be treated as separate validation objectives.

Validation-locked abstention only partially mitigated this problem. Across the evaluated gates, higher accepted-only sensitivity was achieved by reducing coverage, and gate behaviour varied substantially by diagnosis. For the Brazil-adapted model, Mahalanobis retained 79.1% of the combined cohort and 94.9% of the in-scope cohort, yet 74.5% of melanomas remained accepted on the SK/MN route. Across all four scores, only 12.1-18.3% of melanoma SK/MN routes were deferred. Because the thresholds were calibrated exclusively on in-scope validation lesions, unfamiliar diagnoses did not necessarily receive extreme abnormality scores even when their downstream routing was clinically undesirable.

More generally, accepted-only performance is not equivalent to end-to-end safety. Selective-prediction systems should be evaluated jointly by coverage, performance among accepted cases, diagnosis-specific abstention and the residual routing of clinically consequential out-of-scope cases. In the present study, the underlying classifier established the principal sensitivity-specificity operating point, whereas the gates produced smaller within-model changes and did not reliably enforce scope control.

Several limitations should be considered. First, the study was retrospective and included histopathologically confirmed cohorts, so performance in unselected prospective populations remains to be established; the pathology reference standard does, however, fix the labels consistently across all three endpoints and both external sites. Second, the primary diagnostic label space comprised BCC, SK and MN, and the action analysis was a diagnosis-category proxy rather than an observed clinical workflow outcome; the proxy is nonetheless defined from the diagnosis alone and so does not depend on the model being evaluated. Third, the melanoma analysis included 29 lesions, and the thickness endpoint included 26 internal model-only, 24 reader-matched and 21 external lesions, so the corresponding intervals are wide and both endpoints remain exploratory rather than deployment-ready. Fourth, Brazil was available for triage and out-of-scope analyses but not the risk-stratification or thickness endpoints, which restricts the geographic generalisation claim to the triage endpoint. Finally, Mahalanobis was selected for detailed reporting after comparison of four scores rather than prespecified as a confirmatory gate, and all thresholds were calibrated only on in-scope validation lesions. Gate-specific conclusions are therefore exploratory, and the low interception of melanoma SK/MN routes indicates that none of the evaluated gates can be relied on for scope control. Prospective multicentre evaluation should assess broader diagnostic spectra, clinician-AI interaction and actual escalation decisions.

In conclusion, this multi-endpoint BCC-centred framework linked accessible lesion triage, histopathology-informed risk stratification and exploratory dermoscopy-based thickness assessment as complementary pre-biopsy endpoints. Triage macro-AUROC stayed above 0.85 across three cohorts on two continents, performance exceeded the mean of 19 dermatologists on every prespecified primary metric, and the most informative input set differed by endpoint rather than being uniform across them. Matching inputs to the decision at hand is therefore a workable design principle for pathway-level dermatology AI. The adaptation analyses add a second: closed-set performance, out-of-scope routing and selective-prediction performance require separate validation. Prospective multicenter evaluation with clinician interaction is the next step.

## Methods

### Study design and cohorts

This retrospective diagnostic accuracy study included patients with BCC, seborrhoeic keratosis or melanocytic naevus from one development/internal validation source and two external validation cohorts (Figure 1A). The XJTU cohort was assembled at The Second Affiliated Hospital of Xi’an Jiaotong University, Xi’an, China, and contained 1,459 patients diagnosed between December 2020 and April 2025. External validation used the Baoji cohort from Baoji Central Hospital, Baoji, China, for the triage, risk-stratification and thickness endpoints, and the publicly available PAD-UFES-20 Brazilian cohort for the triage endpoint ^25^. The Baoji cohort contained 149 patients contributing 152 lesions, diagnosed between February 2024 and December 2025. The Brazilian three-class cohort contained 846 patients, 1,066 lesions and 1,324 images. Inclusion criteria for the two Chinese cohorts were: (1) histopathological confirmation of BCC, seborrhoeic keratosis or melanocytic naevus; and (2) complete photographic and clinical data. Exclusion criteria were poor image quality, including blurring or obscuration by hair or clothing, or incomplete clinical data (Supplementary Figure S1). Histopathology was the reference standard for diagnostic labels, BCC histopathological risk categories and tumour thickness. Analysis populations are summarized in Supplementary Table S1. The study was approved by the ethics committees of The Second Affiliated Hospital of Xi’an Jiaotong University (2025-215) and Baoji Central Hospital (BZYL2026-65), which waived informed consent because of the retrospective design.

Model-only internal validation and AI-reader comparisons were treated as complementary analyses. AI-reader comparisons used the reader-study analysis set evaluated by dermatologists, whereas model-only validation evaluated algorithm performance on the held-out validation cohorts.

For classification endpoints, XJTU cases were divided at patient level into training and held-out internal test partitions using an approximately 9:1 split, and the development partition was further divided approximately 9:1 into training and validation subsets. The BCC thickness subset was split with stratification by the ≤2 mm label so that thin and thicker lesions were represented in held-out testing. Details are in Supplementary Note S1.

#### Multimodal data collection

In Chinese cohorts, multimodal data included clinical information text, clinical photographs and dermoscopic images. Pathology reports were linked to electronic medical records by patient ID to retrieve diagnostic, histopathological-subtype and tumour-thickness fields. For tumour thickness, if biopsy and subsequent complete excision specimens provided discrepant measurements, the deeper value was used. Clinical text including a structured chief complaint covering lesion location, type, associated symptoms, and disease duration, as well as sex and age, was extracted from electronic medical records. Clinical text examples are shown in Supplementary Note S2. Clinical and dermoscopic images were obtained from hospital skin-image banks and were acquired before biopsy or surgery.

In the XJTU cohort, clinical photographs were acquired using a Canon DS126311 camera or mobile phones, and dermoscopic images were acquired using a BN-PFMF-8001 dermoscope (Nanjing Beining Medical Equipment Co., Ltd.). In the Baoji cohort, clinical images were acquired using a Canon EOS 800D camera, and dermoscopic images were acquired using a DERMOSCOPY-2 dermoscope (Beijing Demete Jiekang Technology Development Co., Ltd.). To reduce confounding from imaging angle, one clinical image per lesion was selected, prioritizing the image taken as perpendicular to the lesion as possible. For BCC lesions, one pre-biopsy dermoscopic image per lesion was used when available.

### Clinical endpoints and primary configurations

We evaluated three staged clinical tasks. Lesion triage was operationalized as BCC versus seborrhoeic keratosis versus melanocytic naevus. BCC risk stratification retained these two common mimics and separated BCC into low-risk and high-risk histopathological categories derived from recorded pathology subtype. The subtype mapping is provided in Supplementary Note S1. Tumour thickness prediction was trained as regression and evaluated as a binary dermoscopy-based decision task in which lesions with measured thickness ≤2 mm were the positive class. The 2 mm cut-off was selected as a management-oriented thin-lesion threshold, as patients with low-risk BCC lesions ≤2 mm in thickness may achieve the highest complete response rates to nonsurgical treatments such as photodynamic therapy or curettage and electrodesiccation ^26–28^.

The input sets were intentionally aligned with the point of care at which each endpoint would plausibly arise. Clinical images and structured text represent information that can be captured during front-line, community or remote assessment. Dermoscopy was treated as a specialist-assessment modality and was evaluated primarily for the thickness endpoint, where the question shifts from lesion recognition towards whether a lesion is thin enough for non-surgical options to be considered. Given the small number of lesions with paired pre-biopsy dermoscopy and measured thickness, this endpoint was prespecified as an exploratory rule-in analysis rather than as a treatment-planning tool.

All clinically plausible modality combinations were evaluated, but the primary result presentation was organized around configurations that matched the intended point of care. The classification analyses emphasized clinical image plus structured text because these inputs are compatible with scalable visual-contextual assessment. Thickness analyses prioritized dermoscopy-based policies to enable high-precision identification of BCC lesions predicted to be ≤2 mm after specialist assessment. Complete modality and threshold policy comparisons are described in Supplementary Note S2.

### Model architecture and training

Clinical and dermoscopic images were encoded using frozen PanDerm ^14^ large-patch16-224 vision encoders (Figure 1B). PanDerm was pretrained by self-supervised learning on 2,149,706 unlabelled images from 11 in-house and public sources. PAD-UFES-20 was not included in this pretraining corpus and was used only as a downstream benchmark in the PanDerm study. The XJTU-trained zero-shot Brazilian estimates were therefore out of sample with respect to both encoder pretraining and task-head training. Brazil-adapted estimates used patient-grouped local adaptation as described in Supplementary Note S5. The out-of-scope analysis excluded every patient used for adaptation training. Structured clinical text was embedded using frozen Qwen3-Embedding-8B ^29^ into 4,096-dimensional vectors, and Qwen itself was not fine-tuned. Text embeddings were projected from 4,096 to 1,024 and then 512 dimensions, with SiLU activation, dropout and layer normalisation. Each available modality token was mapped to a 512-dimensional space and processed by an eight-head self-attention block, residual feed-forward layers and learned modality-weighted pooling. The fused representation passed through a multilayer perceptron and a linear task head, with three or four logits for classification and one scalar output for thickness regression. Classification used cross-entropy loss and thickness regression used Huber loss. Models were optimized with AdamW, 10-step linear warm-up followed by cosine decay, batch size 128 and a maximum of 1,000 epochs. Early stopping used a seed-specific validation split and monitored macro-F1 for classification and mean absolute error for thickness regression. Further implementation details are provided in Supplementary Note S2.

### Dermatologist reader study

Nineteen dermatologists from six tertiary hospitals in China formed the shared reader panel used for all AI-reader comparisons (Figure 1C). Reader-study design and reader characteristics are summarized in Supplementary Note S3 and Supplementary Table S2. Readers evaluated the endpoint-matched XJTU test sets using the information available for each endpoint. The primary comparison used mean reader performance and paired case-bootstrap differences against the AI seed-mean statistic on the same cases.

### Out-of-scope action analysis and abstention gates

The out-of-scope analysis combined the fixed Brazilian BCC/SK/MN test set with ACK, SCC and melanoma after excluding every patient used for Brazilian adaptation training. The exclusion counts and the resulting cohort composition are given in Supplementary Note S7. For each model and seed, three-class probabilities were averaged across images sharing the same patient_id-lesion_id pair before assigning a lesion-level class. BCC, ACK, SCC and melanoma were grouped as warranting further assessment, whereas SK and MN were grouped as non-action. Predictions of BCC therefore indicated further assessment, and predictions of SK or MN indicated the non-action route. This defined the clinical-action direction derived from the three-class output.

We evaluated four lesion-level abnormality scores, MSP^30^, Entropy^30^, Energy^31^, Mahalanobis^32^, with larger values indicating greater abnormality: ^33^

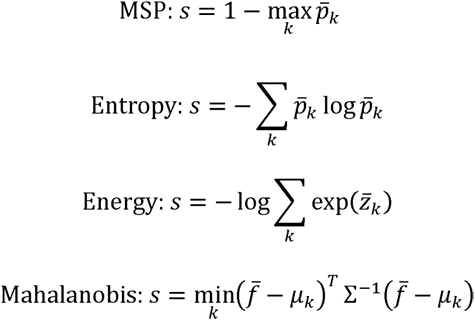

The Mahalanobis score used 512-dimensional lesion-mean features, class centres and a shared Ledoit-Wolf covariance ^34^. For each model, seed and score, the threshold was the 95th percentile of all in-scope validation lesion scores, using the higher quantile rule. Prediction correctness was not used to select the threshold, and no out-of-scope PAD-UFES-20 lesion was used for threshold calibration. Scores strictly above the threshold were abstained. MSP was the reference score. Entropy, energy and Mahalanobis were comparative analyses. Mahalanobis was selected for detailed reporting after the four methods had been compared. Full gate definitions and the per-gate results are given in Supplementary Note S8.

Accepted-only sensitivity and specificity were calculated after excluding abstained lesions and were reported with coverage, defined as the proportion of all lesions retained by the gate. In the system-routing analysis, abstained lesions were assigned to further assessment as an analytical workflow assumption. Gate behaviour was summarized at the validation-locked threshold as the proportion abstained in the in-scope group and separately for ACK, SCC and melanoma. Two summary quantities were computed for each gate: the in-scope abstention rate, and the proportion of melanoma SK/MN routes it deferred. Neither was assessed against an external standard; both are reported as continuous values with intervals.

### Statistical analysis

For the primary endpoints, each metric was calculated separately for the five seed-specific prediction sets and then averaged. Confidence intervals were estimated with 2,000 stratified nonparametric case-bootstrap replicates. For Figures 4-5 and Supplementary Figures S10-S12, 2,000 patient-cluster bootstrap replicates were used so that all lesions and images from the same patient were resampled together. Within each replicate, metrics were recalculated for each seed and then averaged. Seeds were not treated as independent cases. Dermatologist performance was the mean metric across 19 readers. AI-reader comparisons used paired stratified case-bootstrap differences on the same cases. Tests were two-sided and P<0.05 was considered statistically significant.

## Data availability

Individual-level data consisting of clinical and dermoscopic photographs from the XJTU and Baoji cohorts are not publicly available due to patient confidentiality. PAD-UFES-20 is publicly available from its original publication. However, source data underlying all figures and tables, comprising derived numerical values without patient-level images, are accessible from the corresponding author upon reasonable request.

## Code availability

Code for model evaluation, patient-disjoint out-of-scope analysis, abstention gates and figure generation will be released upon publication.

## Author contributions

Z.C., Q.D., S.G., and L.G. conceptualized the work. Z.C. led clinical data curation, coordinated data collection across the XJTU and Baoji cohorts, secured funding and ethics approval, and co-drafted the manuscript. Q.D. conducted all deep-learning experiments, performed data analyses, and drafted the manuscript. B.H. provided technical support for large-scale data processing and contributed to data analyses. Y.G., W.C., W.Z., and L.Z. contributed to clinical data collection and image preprocessing. S.G. provided clinical supervision for the XJTU cohort, while W.C. oversaw clinical supervision for the Baoji cohort. R.E., B.W., and L.G. supervised the technical design of the multimodal framework, the pre-training strategy, and the statistical validation plan. All authors reviewed and approved the final version of the manuscript.

## Acknowledgements

This work was supported by the National Natural Science Foundation of China (82404139), the Shaanxi Province Key Industrial Innovation Chain (2024SF-ZDCYL-01-14), and the Innovation Capability Support Program of Shaanxi Province (2022TD-48).

