## Supplementary materials for "Endpoint-aligned artificial intelligence for biopsy-sparing assessment of suspected basal cell carcinoma"

Supplementary Information

### Abbreviations

ACK, actinic keratosis; AI, artificial intelligence; AUROC, area under the receiver operating characteristic curve; BCC, basal cell carcinoma; CI, confidence interval; CUDA, Compute Unified Device Architecture; F0.5 and F1, weighted harmonic means of precision and recall, weighting precision twice as heavily and equally respectively; Grad-CAM, gradient-weighted class activation mapping; MAE, mean absolute error; MLP, multilayer perceptron; MN, melanocytic naevus; MSP, maximum softmax probability; OOD, out-of-distribution; PAD-UFES-20, the public Brazilian skin-lesion dataset used for external triage validation; RGB, red-green-blue; SCC, squamous cell carcinoma; SiLU, sigmoid linear unit; SK, seborrhoeic keratosis; XJTU, The Second Affiliated Hospital of Xi'an Jiaotong University.

### Supplementary Note S1. Dataset partitioning, endpoint definitions and analysis sets

For the classification endpoints, the XJTU data were split by patient into development and internal test partitions before seed-specific model training. The development partition was then further divided into training and validation subsets. The Baoji and Brazilian cohorts were used as independent validation cohorts according to endpoint availability.

Three analysis settings are used throughout the supplementary tables. The internal test set summarizes model performance on held-out XJTU cohort cases. The reader-study analysis set is the subset evaluated by the dermatologist panel and is used for direct AI-reader comparisons. External validation sets summarize model transfer to the Baoji cohort and, for the 3-class triage endpoint, the Brazilian cohort. Full analysis-set definitions, sample sizes, class distributions, and split descriptions are provided in Table S1.

For BCC risk stratification, recorded BCC pathology subtypes were mapped into histopathological risk categories before model training. Low-risk BCC included nodular, superficial, and fibroepithelioma of Pinkus subtypes. High-risk BCC included micronodular, infiltrative, basosquamous, and sclerosing/morpheaform-type labels. For BCC risk stratification with mimics, the analysis included BCC cases and non-BCC cases retained their seborrhoeic keratosis or melanocytic naevus (MN) labels. For BCC risk stratification without mimics, the analysis was restricted to BCC cases. These analyses assess whether the BCC risk-stratification model retained signal for pathology-defined BCC risk groups after excluding non-BCC diagnostic classes.

For thickness prediction, splitting was performed within the dermoscopy-available BCC subset and stratified by the ≤2 mm binary label so that both thin and thicker lesions were represented in the held-out test set. The ≤2 mm thickness endpoint was defined as a management-oriented thin-lesion threshold. This cut-off is clinically relevant because topical photodynamic therapy has been approved in Europe for selected patients with nonaggressive, low-risk BCC, including superficial BCC and thin nodular BCC with tumour thickness not exceeding 2 mm, when other treatments are impractical or contraindicated^24-26^. The primary operating policy therefore prioritized precision for lesions predicted to be ≤2 mm.

### Supplementary Note S2. AI model performance analysis and bootstrap inference

Encoders and text features. Clinical photographs and dermoscopic images were encoded with PanDerm large-patch16-224 encoders initialised from panderm_ll_data6_checkpoint-499.pth. In the submitted experiments, the image or dermoscopy encoder was frozen, and trainable parameters were limited to modality projections, the attention-fusion block, the final MLP and the task head. Structured text was embedded offline with Qwen3-Embedding-8B into 4096-dimensional vectors, and the Qwen model was not fine-tuned. XJTU and Baoji text embeddings used age, sex, chief complaint and lesion location. PAD-UFES-20 text embeddings used age, sex, lesion location and binary symptom/history variables for itching, recent growth, pain, recent change, bleeding and elevation. PanDerm was pretrained on 11 data sources spanning four imaging modalities, including MYM and HOP (total body photography and dermoscopy), MMT, ACEMID dermatopathology, NSSI, Edu1, Edu2, ISIC2024, TCGA-SKCM and UAH89k. PAD-UFES-20 is listed in the PanDerm study among the downstream evaluation datasets and not among the pretraining sources (ref. 13).

Fusion method. The submitted image-plus-text models used attention fusion. The 4096-dimensional Qwen embedding was first transformed by Linear(4096, 1024), SiLU, Dropout(0.10), Linear(1024, 512) and LayerNorm(512). PanDerm image outputs were 1000-dimensional and were projected to the same 512-dimensional attention space by Linear(1000, 512), LayerNorm(512) and Dropout(0.05). Text tokens entering the fusion block were similarly projected by Linear(512, 512), LayerNorm(512) and Dropout(0.05). A learned modality positional embedding was added for up to four modality tokens.

The modality tokens were processed by a pre-normalised multi-head self-attention layer with embedding dimension 512, 8 attention heads and dropout 0.10. The attention output used a residual connection, LayerNorm(512), a feed-forward block Linear(512, 1024), SiLU, Dropout(0.10), Linear(1024, 512), Dropout(0.10), and a second residual path. Modality tokens were then aggregated by learned softmax pooling weights, followed by LayerNorm(512), Linear(512, 512), LayerNorm(512) and Dropout(0.05). For single-modality models, the same attention-fusion wrapper was retained and operated as a trainable projection/pooling block for the available modality.

Task heads. The fused representation passed through a final MLP: Linear(512, 1024), SiLU, Dropout(0.10), Linear(1024, 512), SiLU and Dropout(0.05). The classification head was a single linear layer Linear(512, C), where C=3 for BCC/SK/MN triage (MN, melanocytic naevus) and C=4 for low-risk BCC/SK/MN/high-risk BCC stratification. The thickness model used a single linear regression head Linear(512, 1). No additional activation function, batch normalisation layer or calibration layer was applied after the task head.

Optimisation and stopping. Classification models were trained with cross-entropy loss and the thickness model with Huber loss (delta=3.0 for the submitted thickness model). All submitted configurations used AdamW with learning rate 5 x 10^-4^ and weight decay 1 x 10^-4^. The learning-rate schedule used 10 warm-up steps with linear increase, followed by cosine annealing over 90% of the maximum epoch budget to a minimum learning rate equal to 1% of the initial learning rate. The batch size was 128, the maximum epoch count was 1000 and early stopping used patience 64 with minimum improvement 0.001. The monitored validation metric was macro-F1 for classification and MAE for thickness regression.

Random seeds. Each endpoint and modality configuration was trained in five seed-specific runs with seed values 42, 43, 44, 45 and 46. The seed controlled the patient-level train/validation allocation, or sample-level allocation when no patient identifier was available, and the corresponding data-sampling path. CUDA deterministic kernels were not enforced. The reported primary estimand is therefore the mean of five seed-specific model fits rather than a claim of exact bitwise reproducibility from a single seed.

Image preprocessing and augmentation. Images were read as RGB. Training, validation and test data used the same deterministic transform: resize the shorter side to 256 pixels, centre crop to 224 × 224, convert to tensor, and normalize with ImageNet mean (0.485, 0.456, 0.406) and standard deviation (0.228, 0.224, 0.225). No stochastic colour or geometric augmentation was used in the formal experiments.

Clinical-text leakage audit. The text input excluded pathology report text, pathology diagnosis, BCC histological subtype, measured tumour thickness and the referring clinician's diagnostic-impression field. Because chief complaint can be entered either as patient-reported symptoms or as clinician-curated free text, we audited the chief-complaint strings in the XJTU and Baoji files for terms corresponding to basal cell carcinoma/BCC, seborrhoeic keratosis, melanocytic naevus, squamous cell carcinoma/SCC, actinic keratosis and melanoma in Chinese, English and Portuguese. No matches were found in the chief-complaint field used for the submitted models.

Clinical-text language and translation. The instruction sentence and field names passed to Qwen were English in all cohorts. XJTU and Baoji field values were kept in the original Chinese electronic-record language for chief complaint, age, sex and lesion location, and no manual or automated translation was performed before embedding generation. The Brazilian/PAD-UFES-20 inputs did not use Portuguese free text. They used English-coded structured metadata fields: age, gender, anatomical region and TRUE/FALSE symptom/history variables.

Example Qwen inputs were as follows. XJTU example: Instruct: Given a patient's chief complaint, age, gender, and lesion location, generate a semantic representation for multimodal skin disease diagnosis. Query: age: 35岁; chief_complaint: 面部丘疹10年; gender: 女; lesion_location: 面部. Baoji example: Instruct: Given a patient's chief complaint, age, gender, and lesion location, generate a semantic representation for multimodal skin disease diagnosis. Query: age: 69岁; chief_complaint: 左眉内侧下方黑色斑疹30年; gender: 男; lesion_location: 左眉内侧下方. Brazilian/PAD-UFES-20 example: Instruct: Given a patient's gender, age, lesion location, skin lesion bleeds, skin lesion changed recently, skin lesion elevated, skin lesion grown recently, skin lesion hurts, and skin lesion itches, generate a semantic representation for multimodal skin disease diagnosis. Query: age: 55; gender: FEMALE; lesion_location: NECK; skin_lesion_bleeds: True; skin_lesion_changed_recently: True; skin_lesion_elevated: True; skin_lesion_grown_recently: True; skin_lesion_hurts: False; skin_lesion_itches: True.

The primary estimand for AI models was the mean performance of five independently trained seeds, used to assess robustness rather than to define a required deployment ensemble. For the primary closed-set endpoints, confidence intervals used 2,000 stratified case-bootstrap replicates. In the formal Brazilian three-class triage analysis, the case and resampling unit was an image, so those intervals do not adjust for multiple images from the same patient or lesion. In main Figures 4-5 and Supplementary Figures S10-S12, confidence intervals instead used 2,000 patient-cluster bootstrap replicates, with all lesions and images from a sampled patient retained together. Metrics were recalculated within each seed before averaging, and seeds were not treated as independent patients.

For classification endpoints, each seed produced a softmax probability distribution over the target classes. Predicted labels were assigned by argmax for accuracy, precision, sensitivity, specificity, and F1-based metrics. Macro-AUROC was computed from seed-specific predicted probabilities and then averaged across seeds. Class-specific AUROC used one-versus-rest targets, whereas multiclass summary AUROC used the macro-averaged multiclass definition implemented in the primary analysis bundle. For each bootstrap replicate, cases were resampled with replacement within outcome strata, and all seed-specific predictions for each selected case were retained. Metrics were recalculated separately for each seed, and the replicate-level statistic was defined as the mean across seeds.

For thickness prediction, the primary decision policy applied each seed’s validation-derived F0.5 threshold unchanged to its corresponding held-out test predictions. Fixed 2 mm and validation-derived Youden policies were retained as sensitivity analyses. For each bootstrap replicate, cases were resampled with replacement within outcome strata, and all seed-specific thresholded predictions were retained. Metrics were recalculated separately for each seed, and the replicate-level statistic was defined as the mean across seeds.

### Supplementary Note S3. Reader-study design and AI-reader comparisons

The primary estimand for dermatologists was the mean performance of the shared 19-reader panel on the reader-study analysis set. The reader panel comprised the 19 dermatologists who completed all three reader-study questionnaires and therefore formed the shared comparator cohort for AI-reader analyses. Readers evaluated endpoint-matched XJTU reader-study cases independently and without access to AI predictions. For lesion triage and BCC risk stratification, readers reviewed clinical photographs with structured clinical text. For thickness prediction, readers reviewed clinical photographs, structured clinical text, and dermoscopy images for the dermoscopy-available subset. For dermatologist performance, each bootstrap replicate resampled cases with replacement within outcome strata and retained all reader decisions for each selected case. Metrics were calculated separately for each reader, and the replicate-level dermatologist statistic was defined as the mean across the 19 readers. AI-versus-dermatologist comparisons used paired stratified case-bootstrap differences on the same cases, comparing the mean across five seed-specific AI predictions with the mean across the 19 dermatologists. Paired case-bootstrap P values are reported for these comparisons.

### Supplementary Note S4. Qualitative Grad-CAM heatmap analysis

Grad-CAM was used for qualitative review of image regions contributing to model outputs on held-out XJTU test cases. For the classification endpoints, class-specific maps were computed from the corresponding output logits. For the tumour-thickness endpoint, the predicted thickness was used as the target output. Gradients were backpropagated through the trained model and endpoint decoder to the image encoder, with the final transformer block norm layer used as the visual target layer. Model weights were not updated during this procedure.

For models that included structured text, the same text embedding used for inference was retained while the image branch was analysed. Patch-level Grad-CAM values were reshaped to the visual-token grid, resized to the input-image resolution and displayed as thresholded colour overlays. Warmer colours denote higher relative saliency within the same image. These heatmaps were reviewed descriptively and were not used for model selection, threshold selection, performance estimation or statistical testing.

### Supplementary Note S5. Brazilian few-shot scaling analysis

For Supplementary Figure S4, PAD-UFES-20 was restricted to BCC, melanocytic naevus and seborrhoeic keratosis. A fixed patient-level test set was selected once and held constant, and contained 132 images from 78 lesions and 27 patients (84 BCC, 24 MN and 24 SK images). Nested training pools targeted 4, 8, 16, 32, 64, 128, 256, 512, 1,024 and 1,192 images while keeping patient groups intact. No patient appeared in both a training pool and the fixed test set. For each target size and modality, the seed-matched XJTU-trained model was the starting checkpoint. PanDerm and Qwen encoders remained frozen, and the attention-fusion module and task head were adapted. Results are five-seed means on the same fixed test set. The smallest pools are representation-probing sensitivity analyses, not evidence that four images are sufficient for deployment.

The 4-image target refers to the labelled Brazilian train-plus-validation pool rather than four independent patients or four observations all used for optimization. Seed-specific patient grouping and validation further reduced the number used for gradient updates. The unadapted XJTU-trained image-plus-text model already achieved accuracy 0.812 (0.768-0.853), macro sensitivity 0.671 (0.594-0.745), macro specificity 0.851 (0.817-0.883) and macro-AUROC 0.905 (0.859-0.942) on the same fixed test set. The smallest points therefore describe local calibration of an existing representation and decision boundary.

### Supplementary Note S6. BCC subclass aggregation control analysis

To test whether the performance difference between the 3-class and 4-class diagnostic settings reflected the added subclass split, we used an external matched-case control restricted to the 136 Baoji cases evaluable by both endpoints. We did not use the XJTU native 3-class and 4-class test partitions for this control because those endpoint-specific internal test sets were not the same cases. For each seed, the 4-class prediction was first assigned by argmax over the original four output probabilities. Predicted low-risk BCC and high-risk BCC were then both mapped to BCC, while seborrhoeic keratosis and melanocytic naevus predictions were retained, and reference labels were collapsed in the same way. Metrics were recomputed for these final hard-label BCC/SK/MN outputs and compared with the dedicated 3-class model on the same 136 Baoji cases.

On the 136 Baoji matched cases, the dedicated 3-class model achieved accuracy 0.825 (95% CI 0.775-0.872), macro-averaged sensitivity 0.839 (0.791-0.882) and macro-averaged specificity 0.917 (0.893-0.939), whereas the 4-class model after hard-decision collapse achieved accuracy 0.882 (0.838-0.922), macro-averaged sensitivity 0.888 (0.844-0.926) and macro-averaged specificity 0.944 (0.923-0.963). Because this control evaluates final predicted classes after the original 4-class argmax decision, probability-based AUROC was not computed for the collapsed output. This external same-case control supports interpreting the 4-class performance decrease mainly as the expected cost of subclassifying BCC risk categories rather than as loss of the underlying BCC/SK/MN distinction.

### Supplementary Note S7. Lesion-level action analysis with adaptation-training patient exclusion

The clinical-action analysis used the fixed in-scope Brazilian BCC/SK/MN test set and ACK, SCC and melanoma records after excluding every patient assigned to the Brazilian adaptation training pool. This removed 130 patients, 175 lesions and 208 images. The retained in-scope set contained 132 images, 78 lesions and 27 patients, and the OOD set contained 766 images, 650 lesions and 532 patients. Their combined set contained 898 images from 728 lesions and 554 patients. Five patients contributed different lesions to both the fixed in-scope and OOD components. The OOD component, rather than the combined set, is therefore described as patient-disjoint from adaptation training.

Within each model and seed, three-class probabilities were averaged across images sharing the same patient_id-lesion_id pair before assigning the lesion-level class. BCC, ACK, SCC and melanoma were assigned to the further-assessment group, and SK/MN to the non-action group. Metrics were calculated separately for five seeds and averaged. Confidence intervals used 2,000 patient-cluster bootstrap replicates. This category mapping defined the clinical-action proxy.

The XJTU-trained 0-shot model had action sensitivity 0.953 (0.940-0.964), specificity 0.579 (0.415-0.729) and balanced accuracy 0.766 (0.683-0.843). The Brazil-adapted model had sensitivity 0.697 (0.668-0.726), specificity 0.979 (0.931-1.000) and balanced accuracy 0.838 (0.811-0.860). Correct melanoma action direction decreased from 0.710 (0.586-0.821) to 0.152 (0.062-0.255), based on 29 lesions from 29 patients (Supplementary Figure S10).

### Supplementary Note S8. Validation-locked abstention-gate analysis

Four abnormality scores were evaluated at lesion level: MSP (one minus the largest lesion-mean class probability), entropy of the lesion-mean probabilities, energy from lesion-mean logits, and squared Mahalanobis distance from the nearest familiar class centre in the 512-dimensional feature space. Higher scores indicated greater abnormality. For every model, seed and method, the threshold was fixed at the 95th percentile of all in-scope validation lesion scores, calculated with the “higher” quantile rule. The threshold did not use prediction correctness or any PAD OOD lesion. MSP was the reference score. Entropy, energy and Mahalanobis were comparative analyses.

Accepted-only metrics exclude abstained lesions and are reported with coverage. System-routing metrics assign an abstained lesion to further assessment as an analytical workflow assumption. Gate behaviour is reported at the validation-locked threshold as the proportion abstained in the in-scope group and separately for ACK, SCC and melanoma. Two summary quantities were computed for each gate: mean in-scope abstention, and the proportion of melanoma SK/MN routes deferred. Both are reported as continuous values; neither was assessed against an external standard.

For the Brazil-adapted model, Mahalanobis was carried forward for detailed reporting as an exploratory post-comparison candidate; its accepted-only metrics are reported together with its lower overall coverage, and no gate was ranked above another. At its locked threshold, it abstained 0.051 (0.009-0.111) of in-scope lesions, 0.266 (0.242-0.290) of ACK lesions, 0.064 (0.035-0.099) of SCC lesions and 0.138 (0.067-0.219) of melanomas. In-scope accepted coverage was 0.949 (0.887-0.990), with accepted-only accuracy 0.973 (0.928-1.000). In the combined action set, accepted-only sensitivity was 0.775 (0.744-0.806), specificity 0.985 (0.948-1.000) and overall coverage 0.791 (0.770-0.813). Mahalanobis abstained 0.121 (0.054-0.202) of melanoma SK/MN routes, and residual accepted SK/MN routing among all melanomas was 0.745 (0.600-0.871). The corresponding diagnosis-specific results for ACK and SCC are shown with melanoma in Supplementary Figure S12 (Main Figure 5, Supplementary Figures S11-S12 and Supplementary Tables S6a-S6c).

### Supplementary tables

Table S1. Analysis populations for the three clinical endpoints. Class abbreviations: SK, seborrhoeic keratosis; MN, melanocytic naevus.

| **Clinical endpoint** | **Analysis set** | **n** | **Class distribution** |
| --- | --- | --- | --- |
| Pre-biopsy lesion triage | XJTU internal test | 146 | BCC, 53; SK, 49; MN, 44 |
| Pre-biopsy lesion triage | XJTU reader-study subset | 146 | BCC, 53; SK, 49; MN, 44 |
| Pre-biopsy lesion triage | Baoji external | 152 | BCC, 54; SK, 46; MN, 52 |
| Pre-biopsy lesion triage | Brazilian external (images) | 1,324 | BCC, 845; SK, 235; MN, 244 (images) |
| BCC risk stratification  with mimics | XJTU internal test | 146 | Low-risk BCC, 25; SK, 49; MN, 44; High-risk BCC, 28 |
| BCC risk stratification with mimics | XJTU reader-study subset | 145 | Low-risk BCC, 25; SK, 49; MN, 44; High-risk BCC, 27 |
| BCC risk stratification with mimics | Baoji external | 136 | Low-risk BCC, 26; SK, 46; MN, 52; High-risk BCC, 12 |
| Tumour thickness ≤2 mm | XJTU internal test | 26 | ≤2 mm, 15; >2 mm, 11 |
| Tumour thickness ≤2 mm | XJTU reader-study subset | 24 | ≤2 mm, 15; >2 mm, 9 |
| Tumour thickness ≤2 mm | Baoji external | 21 | ≤2 mm, 11; >2 mm, 10 |

Table S2. Characteristics of 19 participating dermatologists.

| **Characteristic** | **Value** |
| --- | --- |
| Age, years, median (range) | 36 (29-52) |
| Sex, n (%) | |
| Female | 13 (68) |
| Male | 6 (32) |
| Institution, n (%) | |
| The Second Affiliated Hospital of Xi'an Jiaotong University | 13 (68) |
| External hospitals | 6 (32) |
| Professional title, n (%) | |
| Resident physician | 9 (47) |
| Attending physician | 6 (32) |
| Associate chief physician | 3 (16) |
| Chief physician | 1 (5) |
| Duration of practice, n (%) | |
| ≤5 years | 8 (42) |
| 5-10 years | 5 (26) |
| >=10 years | 6 (32) |

Table S3. Modality sensitivity analyses for the classification endpoints.

| **Clinical endpoint** | **Cohort** | **Input** | **n** | **Macro-F1 (95% CI)** | **Accuracy (95% CI)** | **Sensitivity (95% CI)** | **Specificity (95% CI)** | **AUROC (95% CI)** |
| --- | --- | --- | --- | --- | --- | --- | --- | --- |
| Pre-biopsy lesion triage | XJTU internal test | Clinical image + text | 146 | 0.945 (0.917-0.968) | 0.944 (0.916-0.967) | 0.944 (0.916-0.969) | 0.972 (0.957-0.984) | 0.995 (0.986-0.999) |
| Pre-biopsy lesion triage | XJTU internal test | Clinical image only | 146 | 0.915 (0.878-0.951) | 0.915 (0.878-0.951) | 0.916 (0.878-0.951) | 0.958 (0.939-0.976) | 0.987 (0.973-0.996) |
| Pre-biopsy lesion triage | Baoji external | Clinical image + text | 152 | 0.835 (0.788-0.877) | 0.836 (0.791-0.878) | 0.834 (0.789-0.876) | 0.917 (0.894-0.938) | 0.978 (0.963-0.990) |
| Pre-biopsy lesion triage | Baoji external | Clinical image only | 152 | 0.672 (0.611-0.731) | 0.696 (0.645-0.747) | 0.699 (0.648-0.749) | 0.846 (0.820-0.871) | 0.899 (0.865-0.931) |
| Pre-biopsy lesion triage | Brazilian external | Clinical image + text | 1324 | 0.640 (0.614-0.664) | 0.784 (0.772-0.797) | 0.608 (0.586-0.630) | 0.828 (0.817-0.839) | 0.853 (0.838-0.868) |
| Pre-biopsy lesion triage | Brazilian external | Clinical image only | 1324 | 0.566 (0.540-0.592) | 0.746 (0.733-0.759) | 0.553 (0.531-0.576) | 0.811 (0.800-0.823) | 0.837 (0.824-0.850) |
| BCC risk stratification with mimics | XJTU internal test | Clinical image + text | 146 | 0.780 (0.726-0.830) | 0.829 (0.786-0.870) | 0.783 (0.732-0.834) | 0.946 (0.932-0.959) | 0.943 (0.921-0.964) |
| BCC risk stratification with mimics | XJTU internal test | Clinical image only | 146 | 0.748 (0.691-0.799) | 0.767 (0.715-0.815) | 0.756 (0.701-0.805) | 0.923 (0.906-0.939) | 0.942 (0.919-0.962) |
| BCC risk stratification with mimics | Baoji external | Clinical image + text | 136 | 0.656 (0.598-0.710) | 0.747 (0.696-0.797) | 0.693 (0.627-0.756) | 0.925 (0.910-0.940) | 0.899 (0.863-0.932) |
| BCC risk stratification with mimics | Baoji external | Clinical image only | 136 | 0.489 (0.427-0.551) | 0.525 (0.466-0.588) | 0.542 (0.468-0.617) | 0.854 (0.835-0.874) | 0.823 (0.771-0.875) |

Table S4. Modality sensitivity analysis for the BCC tumour thickness prediction endpoint.

| **Clinical endpoint** | **Cohort** | **Input** | **n** | **Precision (95% CI)** | **Sensitivity (95% CI)** | **F0.5 (95% CI)** |
| --- | --- | --- | --- | --- | --- | --- |
| Tumour thickness ≤2 mm | XJTU internal test | Dermoscopy only | 26 | 0.949 (0.902-1.000) | 0.653 (0.467-0.813) | 0.847 (0.706-0.923) |
| Tumour thickness ≤2 mm | XJTU internal test | Clinical image only | 26 | 0.749 (0.629-0.886) | 0.613 (0.413-0.787) | 0.680 (0.524-0.814) |
| Tumour thickness ≤2 mm | XJTU internal test | Clinical image + text | 26 | 0.812 (0.711-0.915) | 0.667 (0.467-0.827) | 0.748 (0.616-0.849) |
| Tumour thickness ≤2 mm | XJTU internal test | Dermoscopy + text | 26 | 0.893 (0.831-0.956) | 0.547 (0.360-0.720) | 0.766 (0.599-0.862) |
| Tumour thickness ≤2 mm | XJTU internal test | Clinical image + dermoscopy | 26 | 0.798 (0.689-0.917) | 0.680 (0.493-0.827) | 0.732 (0.599-0.846) |
| Tumour thickness ≤2 mm | XJTU internal test | Clinical image + dermoscopy + text | 26 | 0.881 (0.781-1.000) | 0.587 (0.413-0.747) | 0.755 (0.613-0.870) |
| Tumour thickness ≤2 mm | Baoji external | Dermoscopy only | 21 | 0.956 (0.908-1.000) | 0.436 (0.200-0.691) | 0.750 (0.510-0.886) |
| Tumour thickness ≤2 mm | Baoji external | Clinical image only | 21 | 0.828 (0.674-0.971) | 0.545 (0.364-0.727) | 0.719 (0.542-0.848) |
| Tumour thickness ≤2 mm | Baoji external | Clinical image + text | 21 | 0.820 (0.651-0.967) | 0.564 (0.400-0.727) | 0.704 (0.520-0.844) |
| Tumour thickness ≤2 mm | Baoji external | Dermoscopy + text | 21 | 0.819 (0.640-0.969) | 0.527 (0.327-0.727) | 0.705 (0.544-0.850) |
| Tumour thickness ≤2 mm | Baoji external | Clinical image + dermoscopy | 21 | 0.742 (0.510-0.964) | 0.436 (0.236-0.636) | 0.680 (0.452-0.854) |
| Tumour thickness ≤2 mm | Baoji external | Clinical image + dermoscopy + text | 21 | 0.858 (0.646-1.000) | 0.509 (0.309-0.709) | 0.680 (0.529-0.874) |

Table S5. Threshold-policy sensitivity for dermoscopy-only tumour-thickness prediction.

| **Clinical endpoint** | **Cohort** | **Operating threshold** | **n** | **Precision (95% CI)** | **Sensitivity (95% CI)** |
| --- | --- | --- | --- | --- | --- |
| Tumour thickness ≤2 mm | XJTU internal test | Fixed 2 mm cutoff | 26 | 0.795 (0.682-0.906) | 0.827 (0.667-0.947) |
| Tumour thickness ≤2 mm | XJTU internal test | Validation-derived F0.5 | 26 | 0.949 (0.902-1.000) | 0.653 (0.467-0.813) |
| Tumour thickness ≤2 mm | XJTU internal test | Validation-derived Youden | 26 | 0.834 (0.729-0.948) | 0.720 (0.547-0.867) |
| Tumour thickness ≤2 mm | Baoji external | Fixed 2 mm cutoff | 21 | 0.843 (0.749-0.922) | 0.655 (0.455-0.836) |
| Tumour thickness ≤2 mm | Baoji external | Validation-derived F0.5 | 21 | 0.956 (0.908-1.000) | 0.436 (0.200-0.691) |
| Tumour thickness ≤2 mm | Baoji external | Validation-derived Youden | 21 | 0.908 (0.788-1.000) | 0.545 (0.309-0.764) |

Table S6a. Diagnosis-specific abstention at the validation-locked operating point.

| **Gate** | **Role** | **In-scope abstention** | **ACK abstention** | **SCC abstention** | **Melanoma abstention** |
| --- | --- | --- | --- | --- | --- |
| MSP | Reference score | 0.021 (0.002-0.044) | 0.111 (0.096-0.125) | 0.055 (0.026-0.090) | 0.186 (0.110-0.275) |
| Entropy | Comparison | 0.036 (0.005-0.079) | 0.129 (0.113-0.144) | 0.058 (0.026-0.097) | 0.193 (0.109-0.296) |
| Energy | Comparison | 0.059 (0.016-0.122) | 0.165 (0.147-0.182) | 0.053 (0.025-0.083) | 0.152 (0.083-0.233) |
| Mahalanobis | Exploratory post-comparison candidate | 0.051 (0.009-0.111) | 0.266 (0.242-0.290) | 0.064 (0.035-0.099) | 0.138 (0.067-0.219) |

Thresholds targeted 5% validation abstention using in-scope validation lesions only. Values are five-seed mean proportions with 95% confidence intervals from 2,000 patient-cluster bootstrap replicates. Mahalanobis was selected after comparison and was not prespecified. The in-scope set contained 78 lesions from 27 patients.

Table S6b. Accepted-only action performance, coverage and residual melanoma risk.

| **Gate** | **Accepted-only sensitivity** | **Accepted-only specificity** | **Overall coverage** | **Abstained melanoma SK/MN routes** | **Residual accepted melanoma SK/MN routing** |
| --- | --- | --- | --- | --- | --- |
| MSP | 0.723 (0.693-0.753) | 0.978 (0.928-1.000) | 0.904 (0.891-0.917) | 0.183 (0.110-0.267) | 0.703 (0.578-0.816) |
| Entropy | 0.725 (0.694-0.754) | 0.978 (0.928-1.000) | 0.889 (0.874-0.903) | 0.171 (0.093-0.263) | 0.710 (0.576-0.829) |
| Energy | 0.727 (0.697-0.757) | 0.978 (0.928-1.000) | 0.863 (0.848-0.879) | 0.139 (0.072-0.223) | 0.738 (0.600-0.850) |
| Mahalanobis | 0.775 (0.744-0.806) | 0.985 (0.948-1.000) | 0.791 (0.770-0.813) | 0.121 (0.054-0.202) | 0.745 (0.600-0.871) |

Values are five-seed means (95% CI). Accepted-only metrics exclude abstained lesions. Abstained melanoma SK/MN routes are the fraction of no-gate melanoma SK/MN assignments removed by the gate. Residual accepted melanoma SK/MN routing is the fraction of all melanomas still accepted in that route. Results use 2,000 patient-cluster bootstrap replicates, and the combined action set contained 728 lesions from 554 patients.

Table S6c. Gate behaviour on the in-scope test set and out-of-scope ranking ability.

| **Gate** | **Accepted in-scope coverage** | **Accepted-only in-scope accuracy** | **Out-of-scope AUROC** |
| --- | --- | --- | --- |
| MSP | 0.979 (0.956-0.998) | 0.964 (0.908-1.000) | 0.685 (0.607-0.756) |
| Entropy | 0.964 (0.920-0.995) | 0.971 (0.924-1.000) | 0.685 (0.605-0.757) |
| Energy | 0.941 (0.877-0.984) | 0.973 (0.925-1.000) | 0.690 (0.615-0.758) |
| Mahalanobis | 0.949 (0.887-0.990) | 0.973 (0.928-1.000) | 0.751 (0.673-0.817) |

Accepted in-scope coverage and accepted-only in-scope accuracy describe the fixed 78-lesion in-scope test set after gating. Accepted-only accuracy is measured on whatever each gate retains, so it is not comparable across gates at different coverage: a gate that abstains more can raise it if its score preferentially removes difficult lesions. Out-of-scope AUROC ranks in-scope against out-of-scope lesions by score and is threshold-free, so it does not describe behaviour at the locked operating point. Values are five-seed means (95% CI) from 2,000 patient-cluster bootstrap replicates.

### Supplementary figures

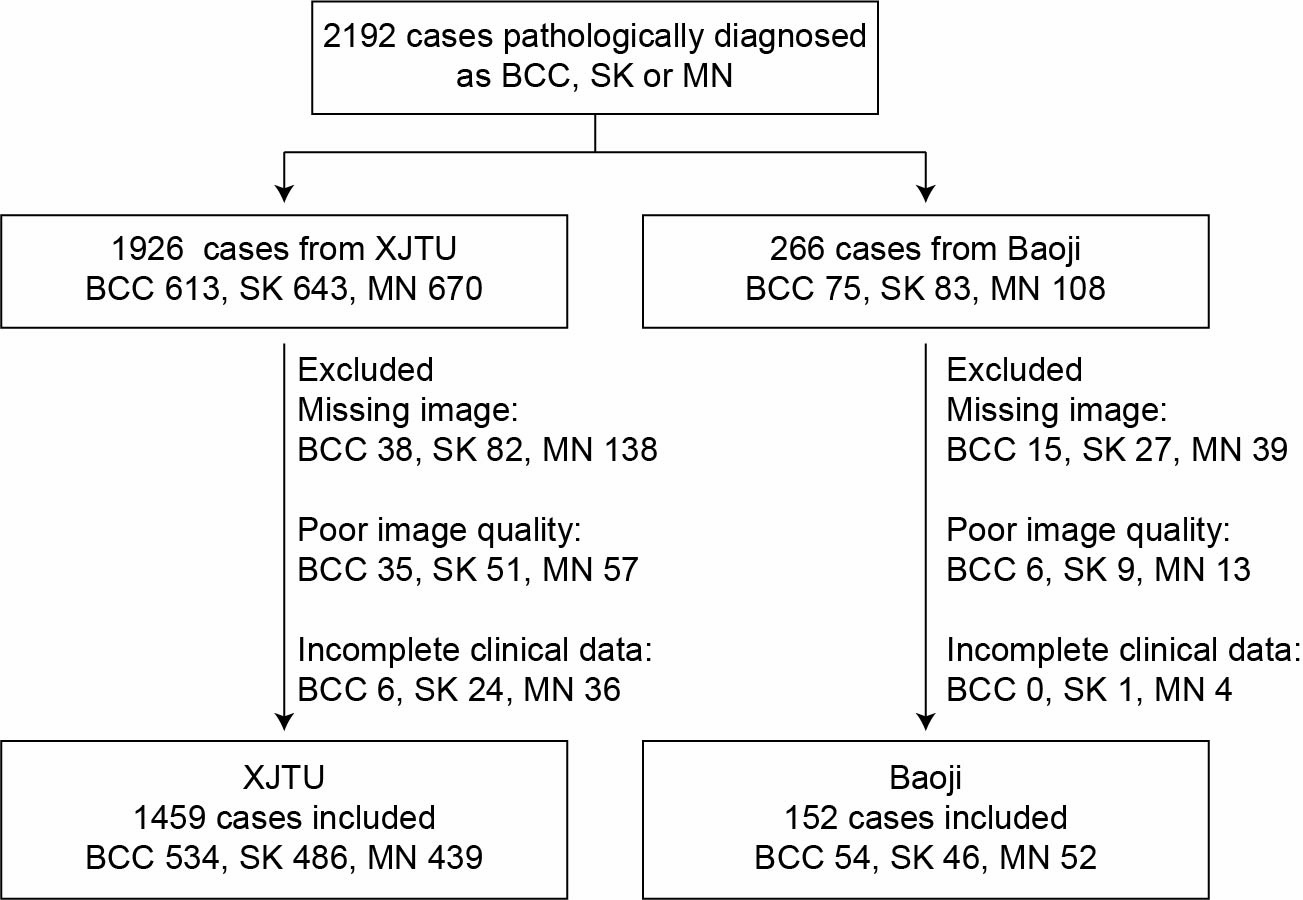

Figure S1. Flowchart of patient screening

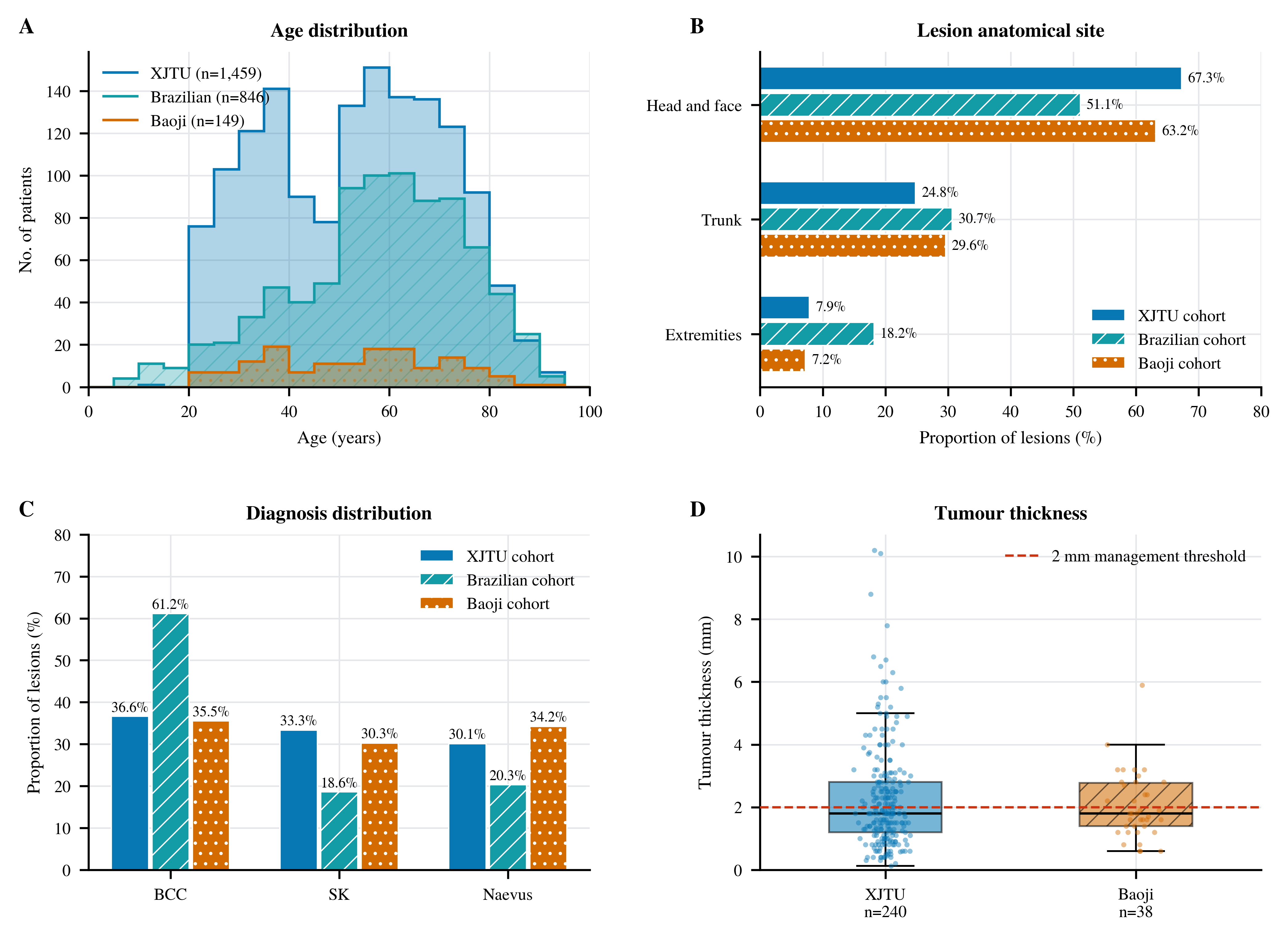

Figure S2. Cohort demographic, lesion-site, diagnosis and tumour-thickness distributions. A, Patient-level age distributions for XJTU (n=1,459), Brazil (n=846) and Baoji (n=149). For Brazil, the median recorded age was used for each patient. B, Lesion-level anatomical-site distributions for XJTU (n=1,459), Brazil (n=1,066) and Baoji (n=152). C, Lesion-level triage-diagnosis distributions for the same cohorts. D, Available histopathological tumour-thickness measurements in the XJTU dermoscopy subset (n=240) and Baoji BCC subset (n=38). These descriptive subsets are distinct from the smaller model-only and reader-matched thickness evaluation sets.

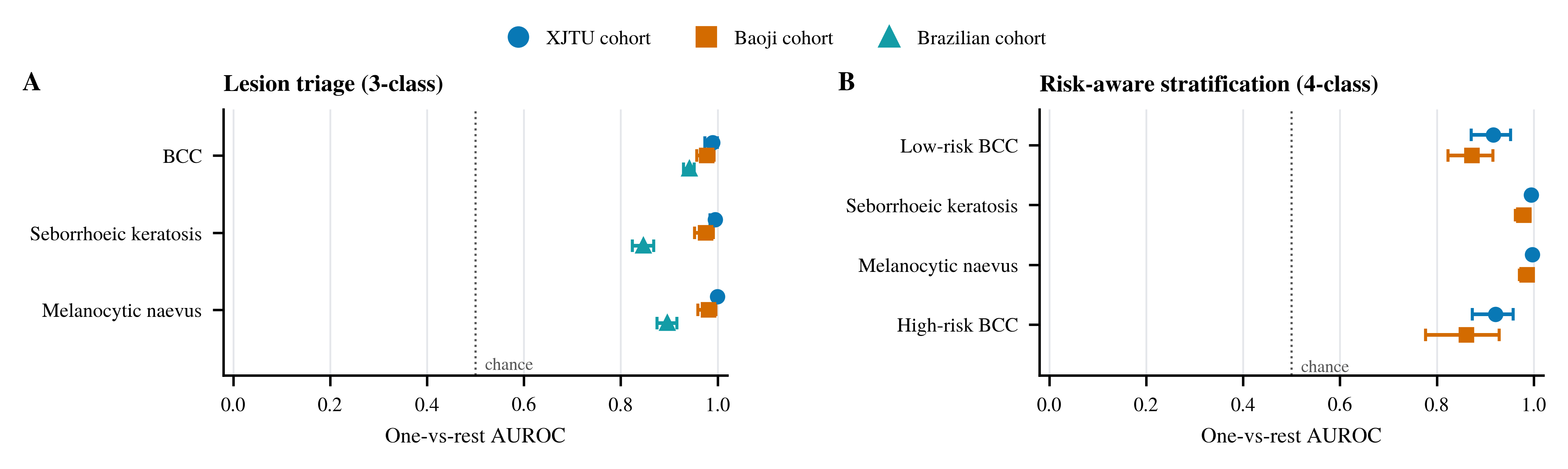

Figure S3. Per-class AUROC for classification endpoints. A, Lesion triage (3-class). B, Risk-aware stratification (4-class). Cohorts are the XJTU, Baoji and Brazilian sets, and the Brazilian cohort contributes to the triage panel only. The dotted rule marks chance (0.5). Points show one-vs-rest AUROC for each diagnostic class, and horizontal intervals show 95% CI from stratified case-level bootstrap resampling. The figure complements the macro-AUROC summaries in the main text by showing which classes contribute to overall discrimination.

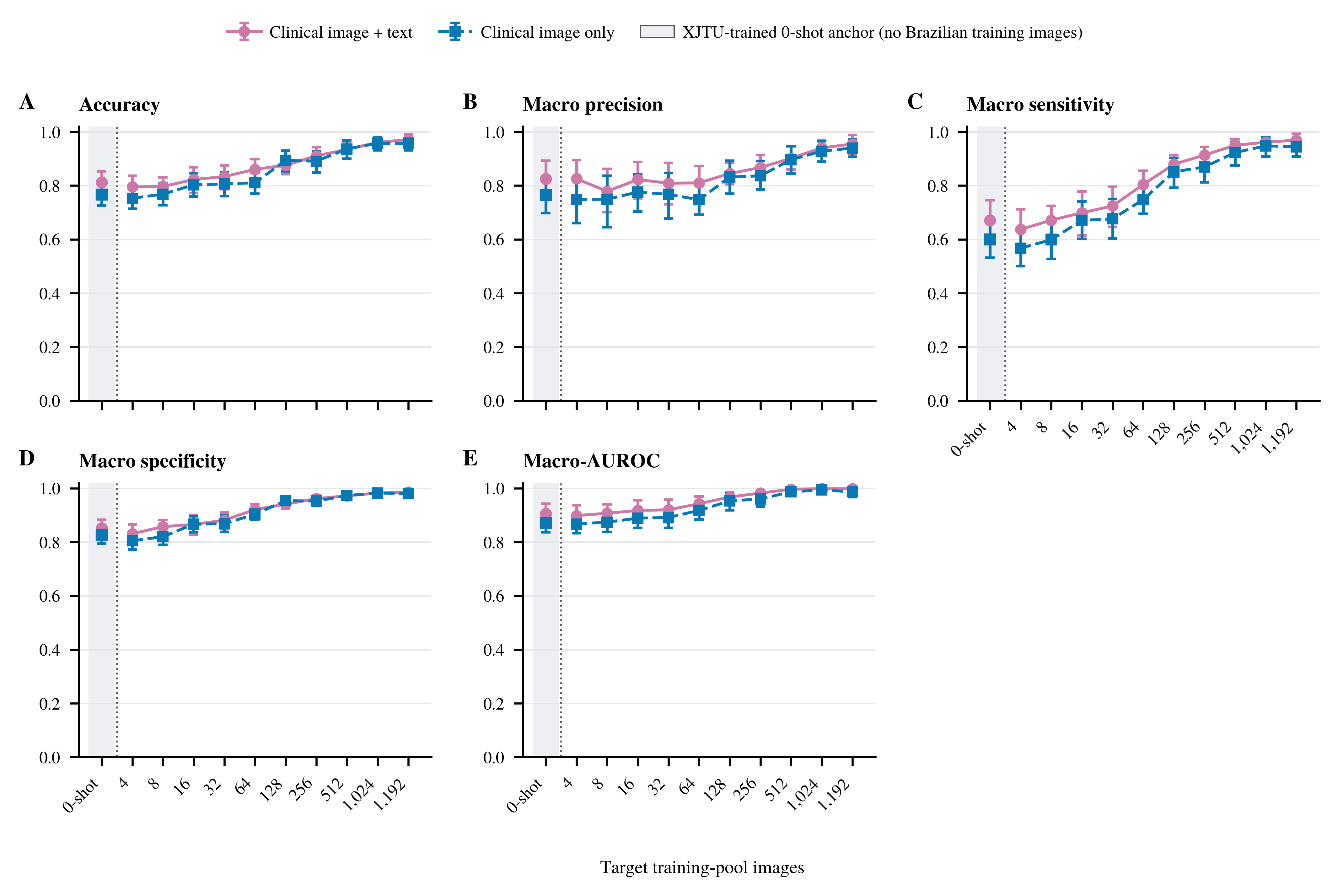

Figure S4. Brazilian cohort few-shot scaling. A, Accuracy. B, Macro precision. C, Macro sensitivity. D, Macro specificity. E, Macro-AUROC. The shaded band and dotted separator mark the unadapted 0-shot anchor. Points show five-seed mean performance for clinical image-only and clinical image plus text models after local fine-tuning on nested patient-grouped PAD-UFES-20 training pools targeting 4, 8, 16, 32, 64, 128, 256, 512, 1,024 and 1,192 images. The 0-shot points show the unadapted XJTU-trained models. Patient groups were kept intact, and the fixed test set was unchanged across all training-pool sizes: 132 images from 78 lesions and 27 patients (84 BCC, 24 MN and 24 SK images). Error bars show 95% image-level case-bootstrap confidence intervals around the five-seed mean and therefore do not adjust for repeated images within patients or lesions. The smallest pools are representation-probing sensitivity analyses, not evidence that four images are sufficient for deployment.

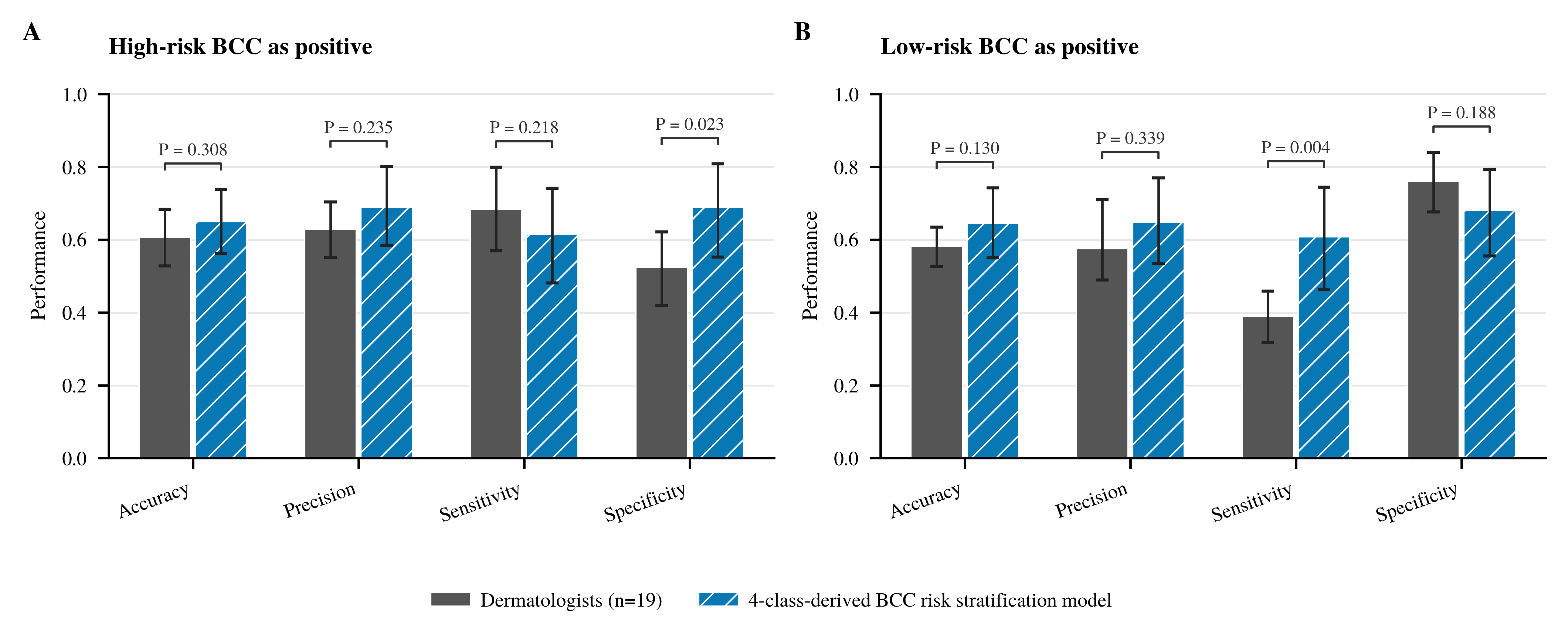

Figure S5. BCC risk stratification without mimics. A, High-risk BCC as the positive class. B, Low-risk BCC as the positive class. The bars compare human readers with the primary clinical image-plus-text model for high- versus low-risk BCC pathological subtyping among BCC cases (BCC-only analysis set, 52 cases). Error bars show 95% CI, and P values are uncorrected two-sided model-versus-reader comparisons.

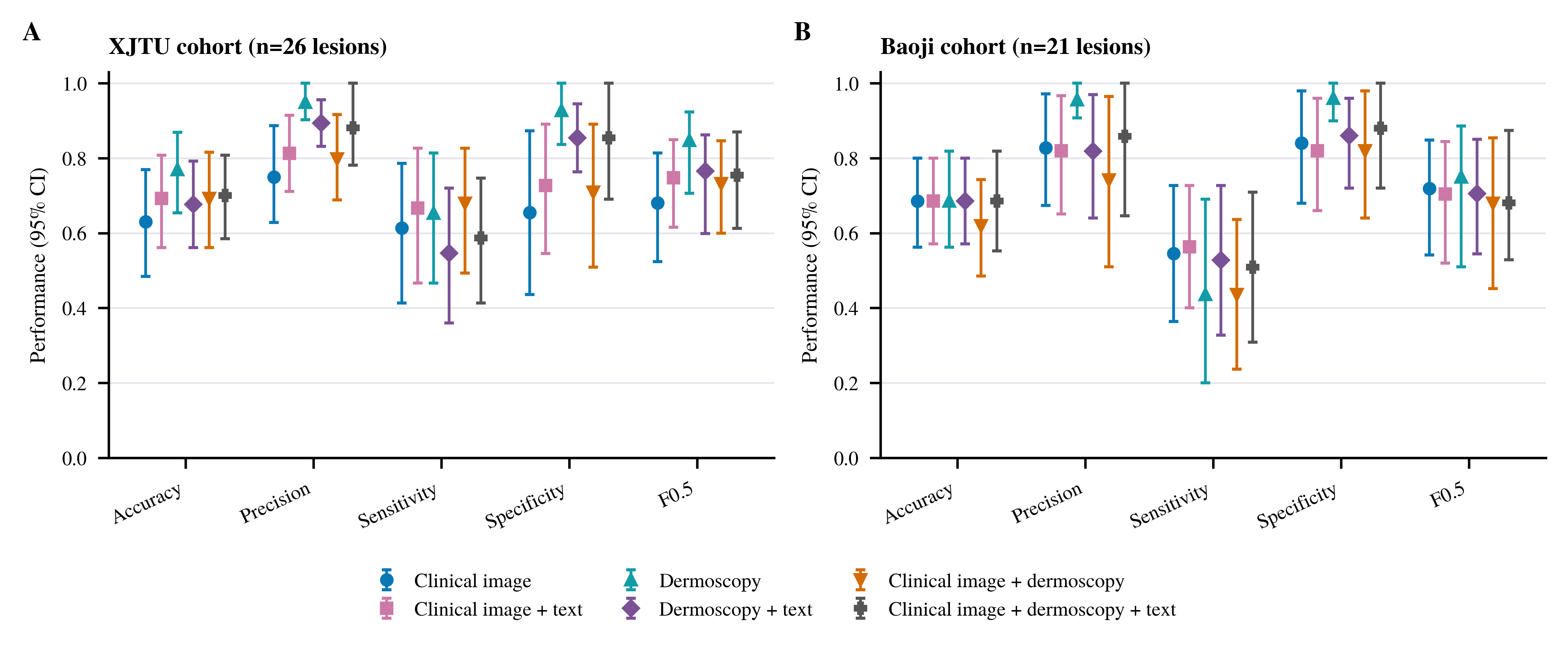

Figure S6. Thickness modality comparison. A, XJTU cohort (26 lesions). B, Baoji cohort (21 lesions). The six series are clinical image, clinical image plus text, dermoscopy, dermoscopy plus text, clinical image plus dermoscopy, and clinical image plus dermoscopy plus text. The plotted F0.5 metric is distinct from the validation-derived F0.5 operating policy under which all series were thresholded. Points show seed-mean performance and 95% CI for thickness ≤2 mm prediction under the validation-derived F0.5 policy. The modality order is consistent across internal and external panels to show endpoint-specific information value.

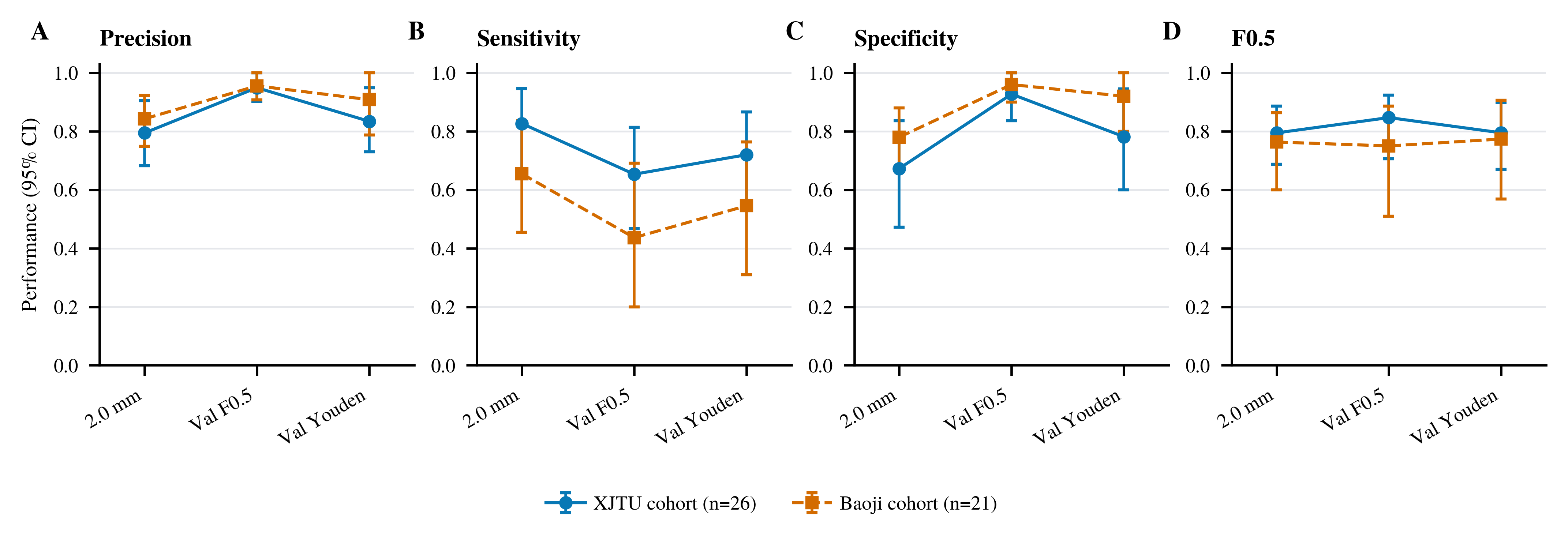

Figure S7. Thickness threshold-policy sensitivity. A, Precision. B, Sensitivity. C, Specificity. D, F0.5. Each panel shows the XJTU (26 lesions) and Baoji (21 lesions) cohorts. Dermoscopy-only performance is shown under fixed 2 mm, validation-derived F0.5 and validation-derived Youden operating policies. This figure shows the trade-off between precision and sensitivity when the regression output is converted to a binary decision, with thickness ≤2 mm as the positive class.

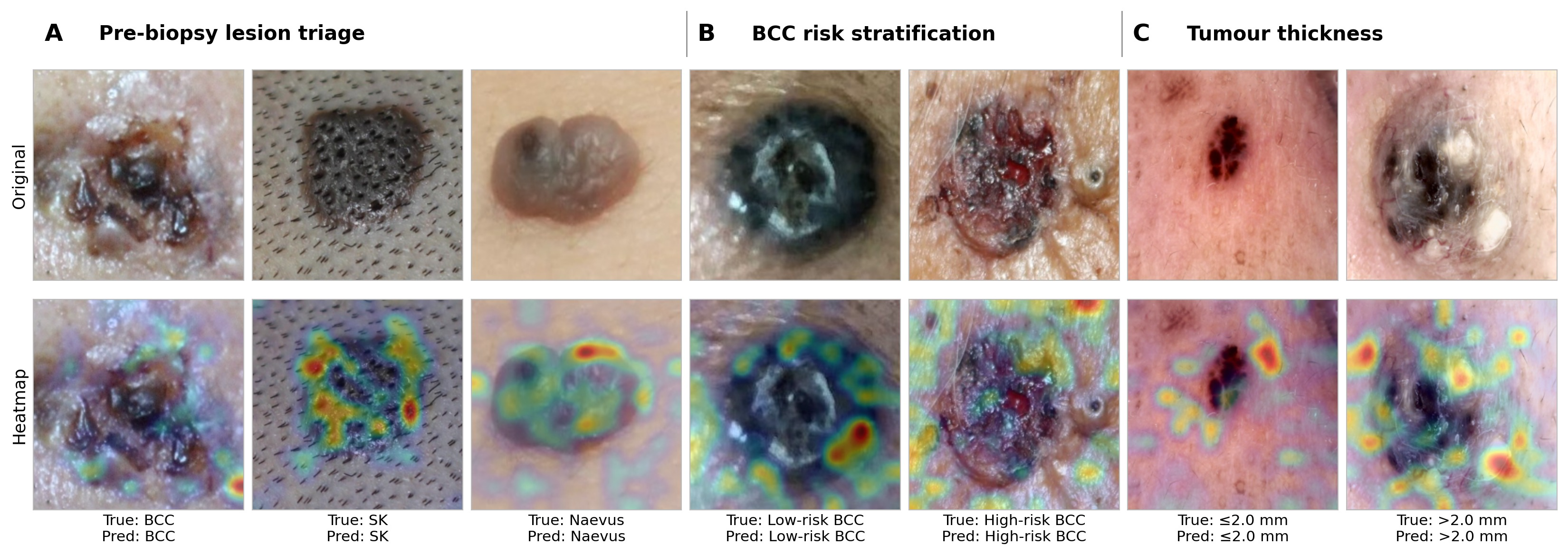

Figure S8. Grad-CAM heatmaps for model interpretation across the three BCC management endpoints.

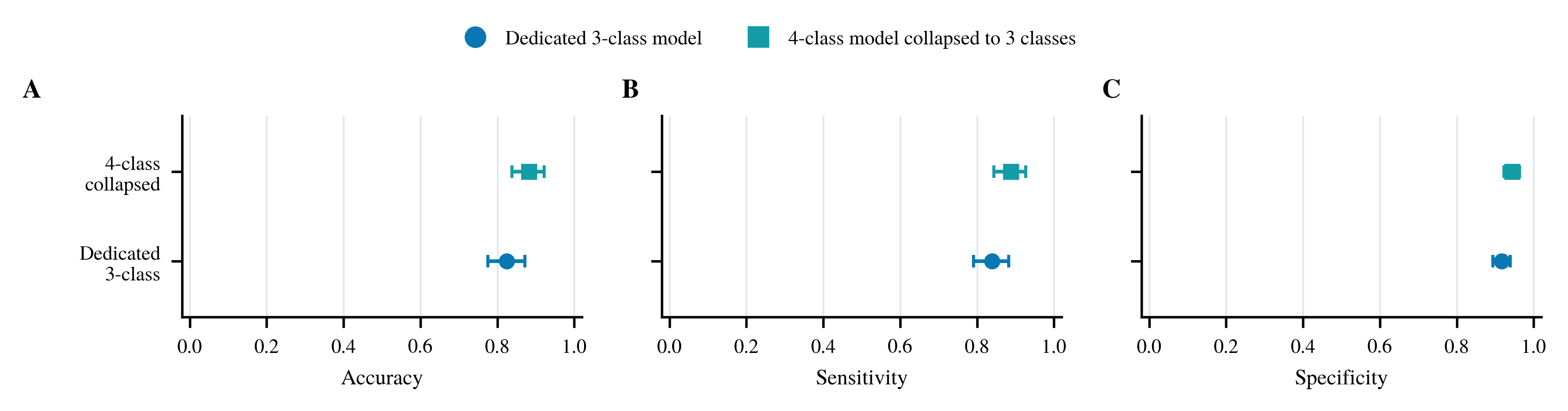

Figure S9. External matched-case control analysis collapsing 4-class BCC subclass decisions to a single BCC class in the Baoji cohort. For each seed, the trained 4-class model was first assigned by its original 4-class argmax decision, and predicted low-risk and high-risk BCC labels were then mapped to BCC, while seborrhoeic keratosis and melanocytic naevus predictions were retained. A, Accuracy. B, Macro-averaged sensitivity. C, Macro-averaged specificity. Points and intervals show seed-mean hard-label performance and 95% case-level bootstrap confidence intervals on the same 136 Baoji cases evaluable by both endpoints, and all panels use full 0-1 axes. The analysis indicates that the apparent 3-class versus 4-class performance gap is driven largely by the added BCC subclass decision rather than failure of the core closed-set BCC/SK/MN distinction.

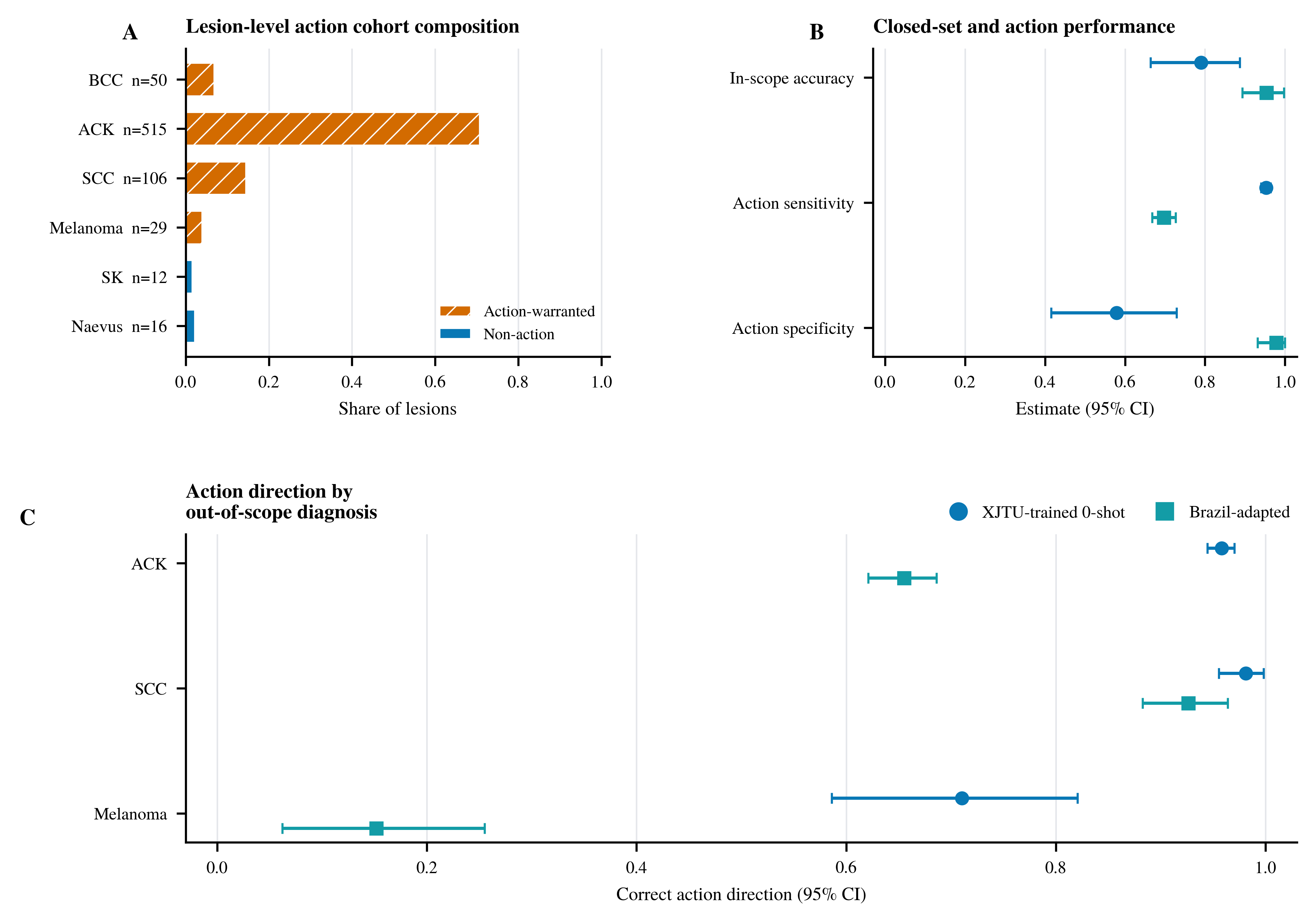

Figure S10. Lesion-level clinical-action analysis after adaptation-training patient exclusion. A, Composition of the 728-lesion combined analysis set. The OOD component excludes all patients used for Brazilian adaptation training. B, In-scope three-class accuracy and overall action sensitivity/specificity for the XJTU-trained 0-shot and Brazil-adapted models. C, Diagnosis-specific correct action direction. Within each seed, probabilities were averaged across images sharing the same patient_id-lesion_id pair. Points are five-seed means and intervals use 2,000 patient-cluster bootstrap replicates.

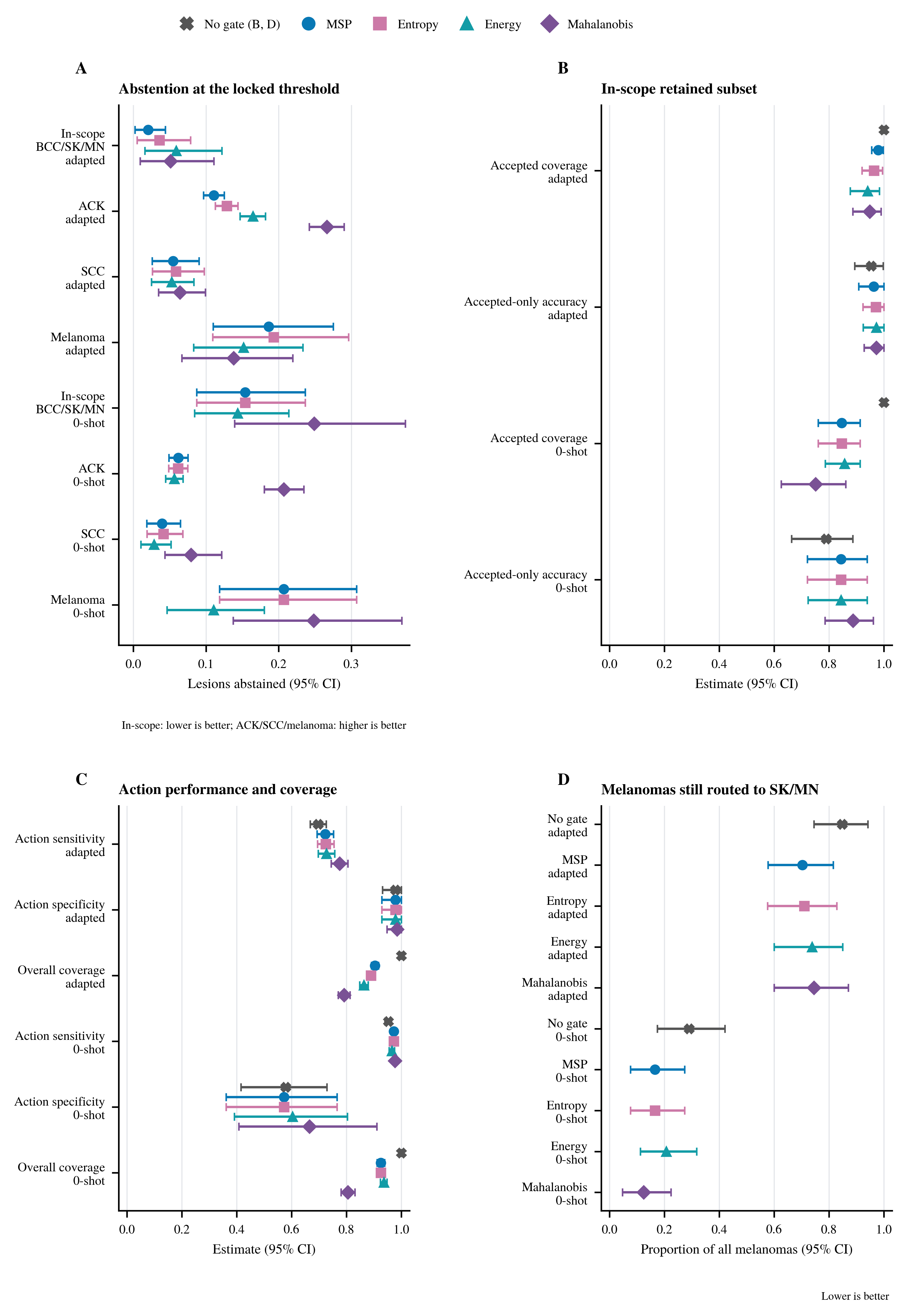

Figure S11. Complete comparison of four validation-locked abstention gates in the XJTU-trained 0-shot and Brazil-adapted models. All four panels show both models, each gate at its own validation-locked threshold for the model it was calibrated on. A, Proportions of in-scope, ACK, SCC and melanoma lesions abstained at the validation-locked thresholds. Lower in-scope abstention indicates fewer familiar lesions deferred, whereas higher diagnosis-specific abstention indicates more out-of-scope lesions deferred. The point estimates run in opposite directions between the models: before adaptation every gate defers more of the familiar in-scope lesions (0.144-0.249) than of the out-of-scope ACK lesions (0.056-0.207), and after adaptation every gate does the reverse (0.021-0.059 against 0.111-0.266). No paired between-model test was performed, so this is a consistent pattern across gates rather than a tested contrast. The 0-shot values apply each gate's own frozen threshold, at the same 5% validation working point, to that model's frozen lesion scores; all sixteen previously published Brazil-adapted values were reproduced before the 0-shot values were computed. B, Accepted coverage and accepted-only accuracy among accepted in-scope lesions, for the Brazil-adapted and XJTU-trained 0-shot models, each with its ungated reference: coverage is 1 without a gate by definition, and the ungated accuracy is the same quantity measured on the full in-scope test set. The unadapted model pays a much larger coverage cost for a lower accepted-only accuracy, 0.751-0.856 coverage at 0.844-0.887 accuracy against 0.941-0.979 at 0.964-0.973 after adaptation. All four gates (MSP, entropy, energy and Mahalanobis) are drawn in every panel. C, Accepted-only action sensitivity and specificity shown with overall coverage, for both models. The two models sit at opposite operating points, ungated 0.953 sensitivity with 0.579 specificity before adaptation against 0.697 with 0.979 after. Those gaps, 25.6 and 40.0 percentage points, are far larger than anything a gate achieves within a model, where the largest shift from the ungated value is 7.8 points of sensitivity in the adapted model and 8.7 points of specificity in the 0-shot model, both under Mahalanobis and both bought with coverage. The model, not the gate, sets the operating point. D, Proportion of all melanoma lesions that remained accepted and were routed to SK/MN without a gate and after each validation-locked gate, for both models. The unadapted model leaves 0.124-0.290 of melanomas on that route and the adapted model 0.703-0.848. The elevated value after adaptation persists across all four gates and without a gate, so it tracks the adapted classifier rather than any particular gate, although the gates do reduce it from 0.848 ungated to 0.703-0.745. Lower values indicate fewer melanomas remaining on the non-action route. Thresholds were fixed from in-scope validation lesions at a target 5% abstention. MSP was the reference score. Mahalanobis was carried forward for detailed reporting only after all four gates had been compared. Points are lesion-level five-seed means and intervals use 2,000 patient-cluster bootstrap replicates.

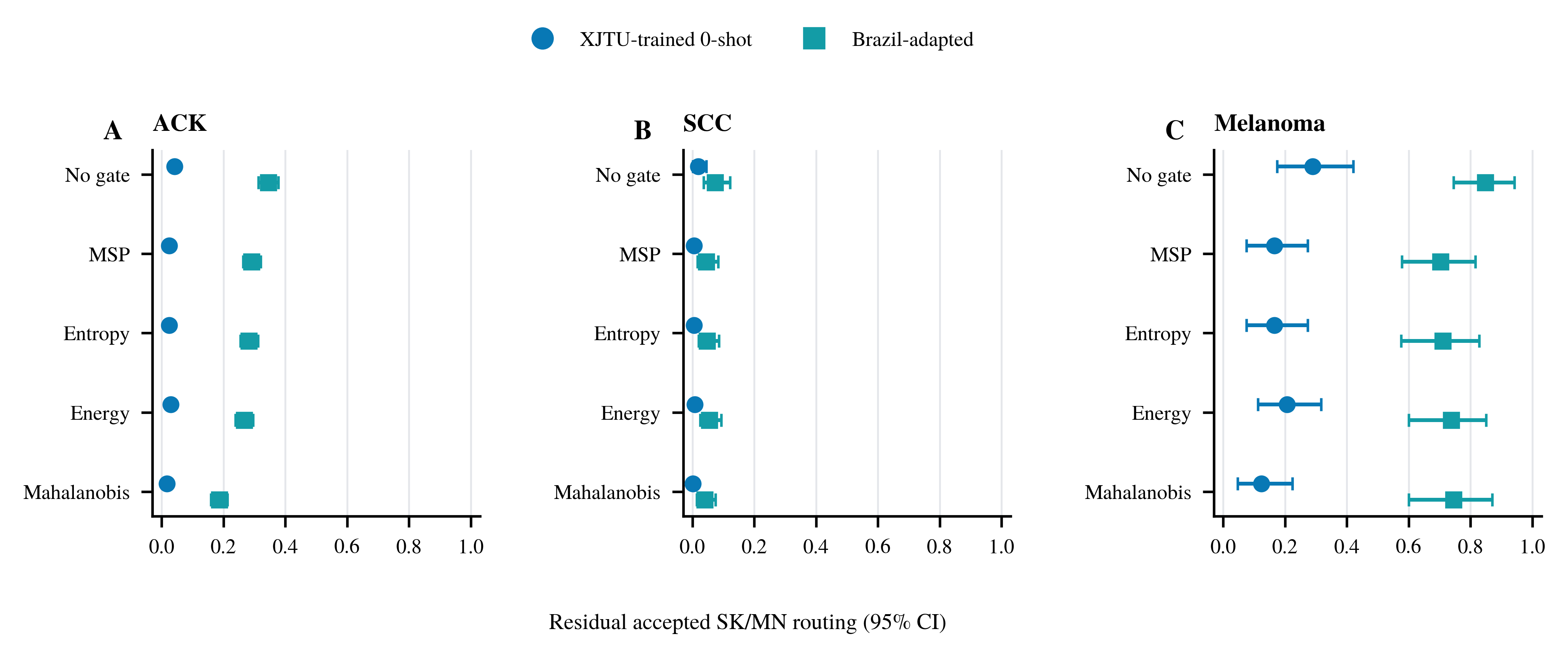

Figure S12. Diagnosis-specific residual accepted SK/MN routing after abstention. A, ACK. B, SCC. C, Melanoma. Within each panel the rows are the ungated reference (No gate) and the four validation-locked scores (MSP, entropy, energy, Mahalanobis), and the two series are the XJTU-trained 0-shot and Brazil-adapted models. Lower values indicate fewer lesions retained in the non-action route. Estimates are lesion-level five-seed means with 2,000 patient-cluster bootstrap replicates, and each estimate uses all lesions of that diagnosis as its denominator.
